# Ignoring spatial heterogeneity in drivers of SARS-CoV-2 transmission in the US will impede sustained elimination

**DOI:** 10.1101/2021.08.09.21261807

**Authors:** Zachary Susswein, Eugenio Valdano, Tobias Brett, Pejman Rohani, Vittoria Colizza, Shweta Bansal

## Abstract

To dissect the transmission dynamics of SARS-CoV-2 in the United States, we integrate parallel streams of high-resolution data on contact, mobility, seasonality, vaccination and seroprevalence within a metapopulation network. We find the COVID-19 pandemic in the US is characterized by a geographically localized mosaic of transmission along an urban-rural gradient, with many outbreaks sustained by between-county transmission. We detect a dynamic tension between the spatial scale of public health interventions and population susceptibility as pre-pandemic contact is resumed. Further, we identify regions rendered particularly at risk from invasion by variants of concern due to spatial connectivity. These findings emphasize the public health importance of accounting for the hierarchy of spatial scales in transmission and the heterogeneous impacts of mobility on the landscape of contagion risk.

## 1 Introduction

Despite widespread transmission of SARS-CoV-2 since late 2019, there has been substantial local variability in implementation of and adherence to public health control measures, both within the United States [1, 2] and around the world [3]. The associated burden of disease has varied by socioeconomic status, with higher infection and mortality rates in marginalized communities [4, 5, 6]. Due to rapid uptake of effective COVID-19 vaccines, case counts in the United States declined through early 2021, but the emergence of SARS-CoV-2 variants – exhibiting higher transmissibility or partial immune escape – and the relaxation of public health control measures have made this a short-lived success story [7, 8, 9, 10].

A central barrier to efficient, targeted control of COVID-19 and the resulting spread of more dangerous variants is the poorly quantified spatial structure of heterogeneity in the determinants of transmission. Local transmission potential (as commonly measured through the time-varying reproductive number, *R*_*t*_ [11]) is a function of a pathogen’s intrinsic transmissibility, local and regional social behavior, and the population immune profile. Heterogeneity in these determinants can be produced by the localised structure of undervaccination, differential rollout and compliance with non-pharmaceutical interventions (NPIs), and socio-economic forces that affect contacts. Contacts provide opportunities for infected individuals to encounter those still susceptible, potentially transmitting COVID-19 with each interaction. Immunity has the opposite effect, with increases in natural and vaccine-induced immunity rates decreasing *R*_*t*_. As social distancing mandates are relaxed – increasing contact rates – the level of population immunity needed for sustained suppression of transmission increases. Similarly, as contact seasonally shifts to indoor settings or mobility seasonally increases, the risk of transmission increases making suppression harder to reach. As a result, the population immunity rate necessary to produce consistent decreases in transmission (i.e. the herd immunity threshold, *R*_*t*_ *<* 1) is variable across time and space, depending on the amount of social behavior in a particular place at a point in time.

Spatio-temporal variation in contact, mobility, and immunity produce heterogeneous transmission potential across time and space. As seen in the case of measles, pockets of low vaccine coverage can enable clusters of sustained local transmission, despite high overall vaccination rates at the state or national level [12, 13]. Similarly, differences in contact rates between communities contribute to heterogeneous transmission rates; sufficient immunity to inhibit sustained transmission for one community’s social behavior may not be enough for another, creating a patchwork of transmission potential. But contagion is also shared across communities, with transmission risk imported and exported through inter-county mobility. Such epidemiological connectivity can substantially complicate the calculus of herd immunity, such that achieving an otherwise sufficient immunity rate in a county may be not be enough to prevent local transmission; at the same time this connectivity can help local interventions (boost in vaccinations, public health control measures) have far-ranging effects beyond the beyond the targeted county. In essence, the epidemiological pictures in neighboring counties contribute to a community’s shared risk of transmission. Yet, yesterday’s transmission landscape is not the same as today’s – natural immunity, vaccination, contact, and mobility all change with time, altering *R*_*t*_. Furthermore, novel variants with potential for higher transmissibility or partial immune evasion will continue to emerge due to low global vaccination coverage, potentially further altering SARS-CoV-2 transmission dynamics [14].

Here, we develop a generative network model to estimate spatially and temporally dynamic transmission potential and parameterize it by pooling empirical data from diverse sources. We estimate mobility through mobile app-based location data, contact rates with a large-scale social survey and a novel metric capturing seasonality in indoor interactions, vaccination through individual state records, and model seroprevalence with state-level studies and county-level incidence data. We demonstrate the roles of within-community contact versus between-community mobility in transmission risk, the role of natural versus vaccine-induced immunity in structuring the susceptibility landscape, the variable impact of potential variant mutations on disease dynamics, and the influence of altering each of these mechanisms in the effectiveness of public health interventions. Our findings highlight the importance of considering disease dynamics at fine spatial scales and the heterogeneous impact of mobility on the landscape of COVID-19 transmission potential, and will help in pandemic response planning in coming months.

## 2 Results

Across the United States, spatial heterogeneity in county-level COVID-19 transmission rates has persisted over the course of the pandemic. There is substantial heterogeneity not just between states, but also within states. We seek to parsimoniously capture this heterogeneity by estimating *R*_*ij*_(*t*), the transmission events generated in community *j* by a case from community *i* at time *t*, based on rich data on contact, mobility, vaccination and seroprevalence in near real-time and at a fine geographic scale. We quantify county-specific transmission potential by aggregating *R*_*ij*_(*t*) in two ways: 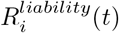 is the potential for cases in a community *i* to generate new cases elsewhere; while 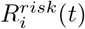 captures the combined potential for new cases within a community *i* generated by transmission from both local cases within the community and regional cases in connected locations, and is analogous to *R*_*t*_ which is traditionally measured from case data [11]. Some of the heterogeneity in transmission potential is systematic, with urban areas exhibiting patterns of transmission distinct from those of suburban or rural counties, but much of it is also unique to the dynamics of specific communities (Figure 1A). In addition to spatial variability, this transmission heterogeneity is itself variable across time. During the summer of 2020, COVID-19 transmission in the US was largely confined to hotspots, with most other areas experiencing only sub-critical transmission. During the late fall and winter, in contrast, COVID-19 transmission was widespread and diffuse. Most counties were experiencing exponential growth in cases. In the spring of 2021, transmission becomes again more focal but distributed differently than before – driven more by patterns of immunity, especially vaccination rates, than by patterns of mobility or contact (Figure 1A).

**Figure 1:**
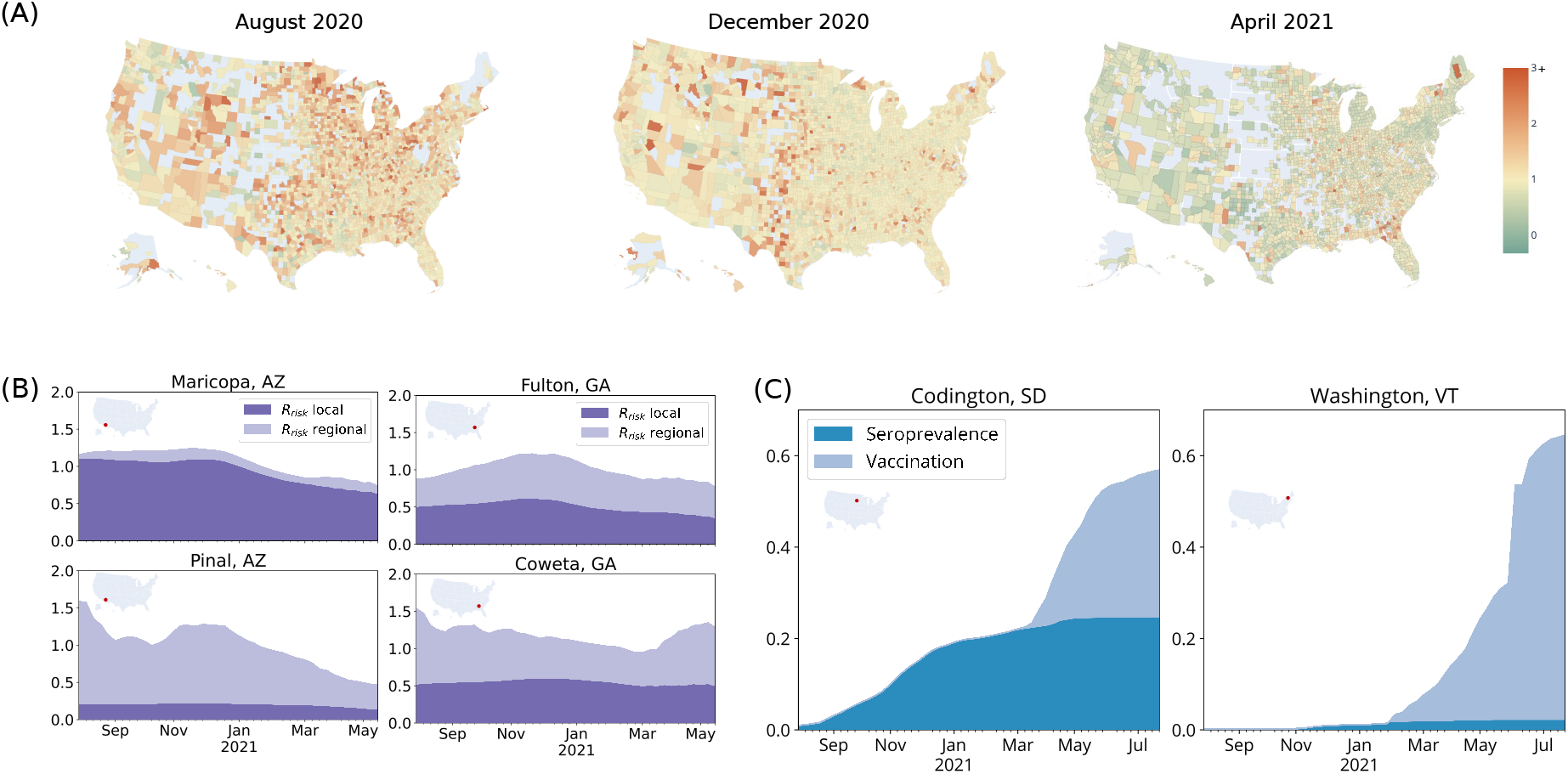
Transmission potential is heterogeneous and driven by contact, mobility and immunity landscapes. (A) *R*_*risk*_ values for three time periods show that transmission potential is spatiotemporally heterogeneous. (Counties in grey are in regions of low incidence and thus *R*_*risk*_ is not defined). (B) We highlight the role of within-community contact versus between-community mobility by considering the local and global components of *R*_*risk*_ in two urban-suburban locations. There is high mobility from Pinal County to neighboring Maricopa County leading to a significant proportion of Pinal’s *R*_*risk*_ coming from the regional component. On the other hand, neighboring Fulton County and Coweta County are less strongly linked through mobility so are more equally affected by the local and regional components of *R*_*risk*_. (C) *R*_*risk*_ is also shaped by the structure of immunity, which evolves over time. Population immunity can be built through infections, causing substantial morbidity and mortality (as in Codington County, South Dakota), or through vaccination (as in Washington County, Vermont).

Realized within-county transmission rates are a function of both intra-county transmission (the local contribution of *R*^*risk*^ driven by within-community contact) and inter-county transmission (the regional contribution of *R*^*risk*^ from mobility-linked communities) (Figure 1B). Transmission potential within counties that act as sink locations is dominated by regional contributions (i.e. absorbing new infections because of significant recurring mobility by residents to other locations). On the other hand, within counties that act as source locations transmission dynamics are driven by local, rather than regional, contributions (i.e. producing new infections because of significant within-county contact and substantially less mobility by residents to other locations). This dynamic is evident in Arizona’s more suburban Pinal County and more urban Maricopa County (which includes the city of Phoenix). Most of Pinal County’s transmission is due to imported risk from outside of the county (i.e. a sink location), while almost all of Maricopa County’s estimated transmission is due to intra-county contact and mobility (i.e. a source location). However, when mobility between a pair of neighboring communities is reasonably symmetric – as is the case in Georgia’s more suburban Coweta County and urban Fulton County (containing the city of Atlanta) – transmission potential is more evenly distributed between the local and regional components for both locations. These differences illustrate the practical importance of taking into account systematic spatial structure as well as the impact of local disease dynamics.

In addition to contact and mobility, local transmission rates are determined by a community’s susceptibility; a community’s potential paths to high rates of immunity can produce dramatically different public health burdens (Figure 1C). Communities like Codington, South Dakota have experienced substantial COVID-19 transmission, leaving approximately 24% of the residents with protection through natural immunity, but have only reached low vaccination rates (with 33% protected through vaccination as of July 25th). On the other hand, the community of Washington, Vermont, has thus far accrued population immunity primarily through vaccination (with 63% of the residents protected through vaccination and 2% protected through natural immunity as of July 25th). The two settings may thus currently have a similar proportion of the population that is fully susceptible to future SARS-CoV-2 transmission, but the path of the South Dakota community comes at the grave cost of 3 deaths for every 1000 residents of the community, and the protection provided by natural infection has is inferior to that of vaccination [15].

The local heterogeneities in mobility, contact, and susceptibility can systematically structure the network of transmission potential (Figure 2A). We find that *R*^*risk*^ – the expected number of secondary cases per primary case in the county – is, on average, lower and more tightly clustered around the mean in urban counties than in rural counties. However, the *R*^*liability*^ – the expected number of secondary infections anywhere deriving from a primary case in the county – is, on average, higher in urban counties than in rural counties. In other words, mobility into core urban locations drives widely distributed networks of transmission in the surrounding suburban and rural locations (see Figure **??**).

**Figure 2:**
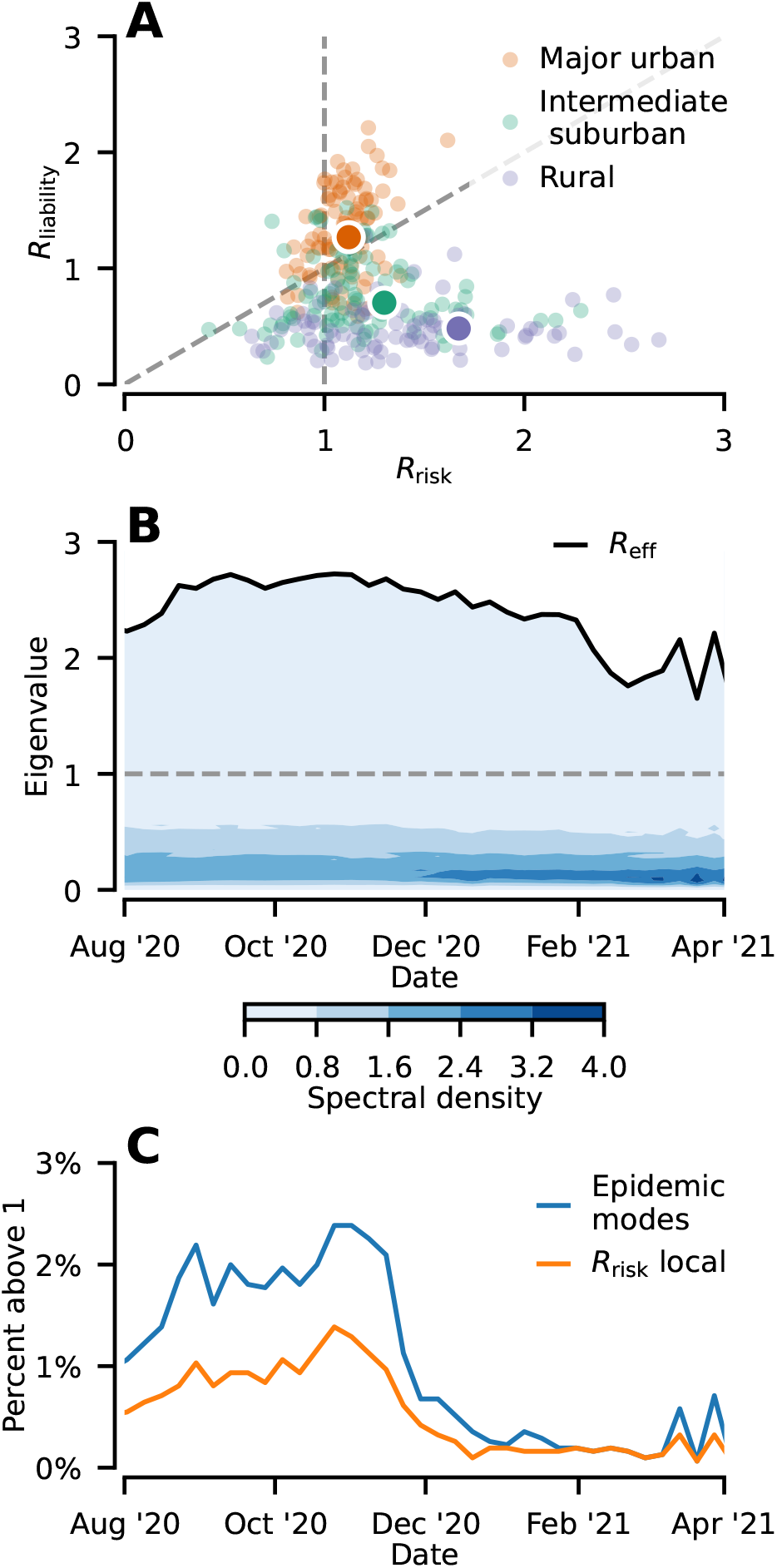
COVID-19 spread is driven by connections between communities. (A) The values of a location’s *R*^*risk*^ and *R*^*liability*^ vary across an urban-rural gradient. Urban locations (green) tend to have higher *R*^*liability*^ and *R*^*risk*^ in a narrow band close to 1. Rural locations (purple) have lowest *R*^*liability*^ and the greatest range in *R*^*risk*^. Intermediate suburban locations (orange) interpolate between the two.(B) The eigenvalue spectrum density of the *R*_*ij*_ matrix over time (blue shading) with the global effective reproductive number, *R*_eff_, highlighted (black line). Each eigenvalue at a given time *t, λ*_*t*_, corresponds to a distinct epidemic mode that either grows (*λ*_*t*_ *>* 1) or shrinks (*λ*_*t*_ *<* 1) in unison. (C) The number of growing epidemic modes (blue) and locations with the local component of *R*^*risk*^ greater than one (orange) through time. The number of growing epidemic modes is consistently greater than the local component of *R*^*risk*^, indicative of extensive betweeen-county transmission.

Our network model integrates the full spatial dynamics of transmission potential. As such, the spectral properties of its adjacency matrix *R*_*ij*_(*t*) give additional insight into the spatial dynamics of the epidemic across time (Figure 2B). The largest eigenvalue of *R*_*ij*_(*t*) corresponds to the global effective reproductive number which, when less than one, indicates that disease transmission has been successfully controlled across the entire country. This corresponds to the epidemic threshold of the next-generation matrix [16, 17]. Although the global effective reproductive number remains roughly constant before the initiation of mass vaccination in early 2021, its consistency obscures substantial spatial variability in transmission. We examine this spatial heterogeneity using the distribution of time-varying eigenvalues of the matrix *R*_*ij*_(*t*), with each eigenvalue corresponding to a distinct epidemic mode. Epidemic modes represent distinct spatially-extensive components of the epidemic dynamics that evolve at the same pace. The magnitude of each mode’s corresponding eigenvalue represents the growth rate of the mode, with a rate greater than 1 indicating growth and less than 1 indicating contraction. The number of eigenvalues greater than 1 gradually increases through the summer and fall, peaking at around 70 distinct growing epidemic modes (roughly 2% of the total) (Figure 2C). This increase indicates that the fall transmission wave in the United States did not occur through one unified surge, but through a number of unique, spatially distinct surges. After December 2020, the number of growing modes abruptly fell, indicating that the remaining case growth fell into a few spatially distinct patterns. Crucially, the number of epidemic modes is always larger than the number of counties with a local component of *R*^*risk*^ larger than one. This relationship indicates that the spatial growth of the epidemic is driven by connected communities, and not solely determined a small number of isolated localities with super-critical transmission. Consideration of local transmission alone would systematically underestimate the kinetics of epidemic growth.

In addition to the contribution of between-county transmission to case growth, we examine the local and regional impact of a county’s public health interventions on transmission control (Figure 3). We present a summary quantity *φ*_*ij*_ that provides an estimate of the increase in vaccination in county *i* required to achieve an equivalent reduction in 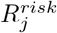 from decreases in within-community (non-household) contacts in county *j* (Figure 3). In other words, a decrease in a county’s average number of contacts by one would be expected to decrease transmission within that county by about as much as an increase in vaccination by *φ*_*ii*_. The distribution of *φ*_*ii*_ values (i.e. within-county transmission reduction) has changed throughout the pandemic (Figure 3A). In the spring of 2020, social distancing was at a high and *φ*_*ii*_ values peaked. Because social distancing interventions decreased contacts, the marginal impact of any further reduction in contact rates was especially high. During the spring of 2021, on the other hand, vaccination rates were rapidly increasing (and social distancing mandates were relaxed), decreasing *φ*_*ii*_. Higher vaccination rates meant that there were more immune individuals in the population, decreasing the infection risk from any particular contact and, in concert, higher overall contact rates decreased the marginal impact of reductions in contact. Consequently, social distancing early in a pandemic is especially effective at curbing spread, buying time to safely build immunity through vaccination. Social distancing later in the pandemic, when immunity is higher, requires even more zealous adherence to achieve the same reduction in transmission. This decrease in the effectiveness of social distancing measures combined with the potential for response fatigue later in the pandemic adds to the compelling evidence for implementing control measures earlier [18]. Additionally, this result suggests that social distancing measures in areas with low vaccination rates may be needed even in case of low epidemic circulation to give time to pre-emptively increase immunity through vaccination.

**Figure 3:**
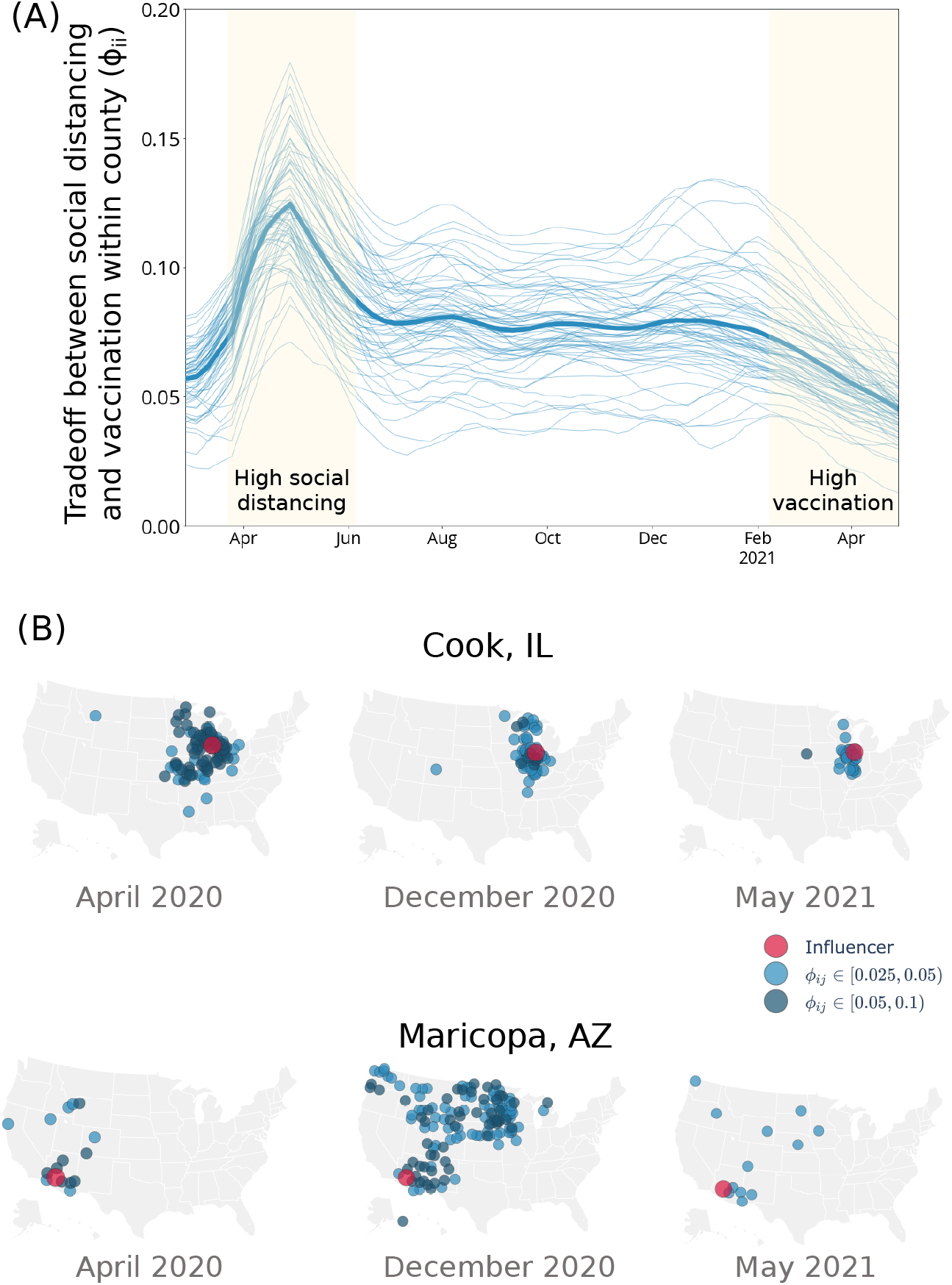
Local and regional impact of public health interventions on transmission control. We consider *φ*_*ij*_ in urban counties: a decrease of the average number of contacts by one in county *j* would be expected to decrease transmission in county *i* by about as much as an increase in vaccination in county *i* of *φ*_*ij*_. (A) The tradeoff between social distancing and vaccination within a given county (measured by *φ*_*ii*_) varies over time. When contact is low (during spring 2020) due to social distancing, every unit reduction in contact amounts to a larger decrease in susceptibility. When vaccination is high, and social distancing is diminishing (during spring 2021), more contact reduction is required to achieve the same gains in vaccination. (B) The public health actions taken by a community can have far reaching consequences. Reductions of contact in Cook County, Illinois effectively translate to reductions in susceptibility across the Midwest, and the influence of social distancing in Maricopa County, Arizona on other communities varies through the pandemic.

In addition to the impact of a county’s social distancing on within-county transmission, we examine the impact of a county’s social distancing on transmission in other counties *φ*_*ij*_; reductions in contact rates in Cook County, IL, and Maricopa County, AZ reduce SARS-CoV-2 transmission throughout the region (Figure 3B). The influence of these urban counties on transmission reductions is heterogeneous across time and space, depending on within-county contact and immunity as well as between-county mobility. In Cook County, the network of transmission reduction is largest in April, 2020 and reduces over time. In Maricopa County, the network of transmission reduction is more focally clustered in April, 2020 before expanding over much of the mid- and north-west in mid-December. In both cases, the network of transmission reduction is smallest in May 2021, reflecting the decrease in *φ*_*ij*_ caused by rising vaccination and decreased social distancing. Although mobility into these urban counties can generate widely distributed networks of transmission, mobility also means that social distancing in these counties have wide-ranging effects, reducing transmission over entire regions.

The benefit of a mechanistic approach to transmission potential estimation is that it allows for scenario planning under assumptions on future interventions, vaccination campaigns, and variant emergence [19]. To understand the impact of each scenario on outbreak risk, we estimate the connected network of communities that have the potential for *R*^*risk*^ ≥ 1, or the set of communities that could experience a simultaneous surge in transmission in the given scenario (Figure 4). After cases rise in this set of identified communities, this surge would likely continue to spread outward to new counties. Given current vaccination levels (average US complete vaccination is at ∼ 49% as of July 28, 2021) and assuming there had been no introduction of variants, almost all COVID-19 transmission in the United States would be sub-critical with current levels of contact and mobility (Fig. 4A). Instead, with the Delta variant, even when we reach a 70% average vaccination rate (with existing vaccination disparities), we would still expect significant transmission clusters (Fig. 4B). A highly effective vaccination campaign in the US, alongside current protective behaviors, is preventing a simultaneous surge in transmission across much of the country, but more work is needed to reach undervaccinated communities, particularly in light of variant transmission. Existing or future variants of concern present spatially heterogeneous risks through a variety of mechanisms. To better understand the role of these mechanisms, we consider the landscape of transmission potential under high variant transmissibility and capability for immune evasion while assuming current levels of susceptibility, contact and mobility. Elevated transmissibility (∼ 100% increase over wild type, similar to the Delta variant) leads to a rise in cases across the majority of counties (Fig. 4C), while immune evasion (∼25% decrease in natural resistance/vaccine effectiveness) leads to several disconnected networks in different regions of the country (Fig. 4D). These scenarios indicate that while vaccination is suppressing transmission, rates of immunity are still low enough that elevated transmissibility enables outbreaks better than immune evasion.

**Figure 4:**
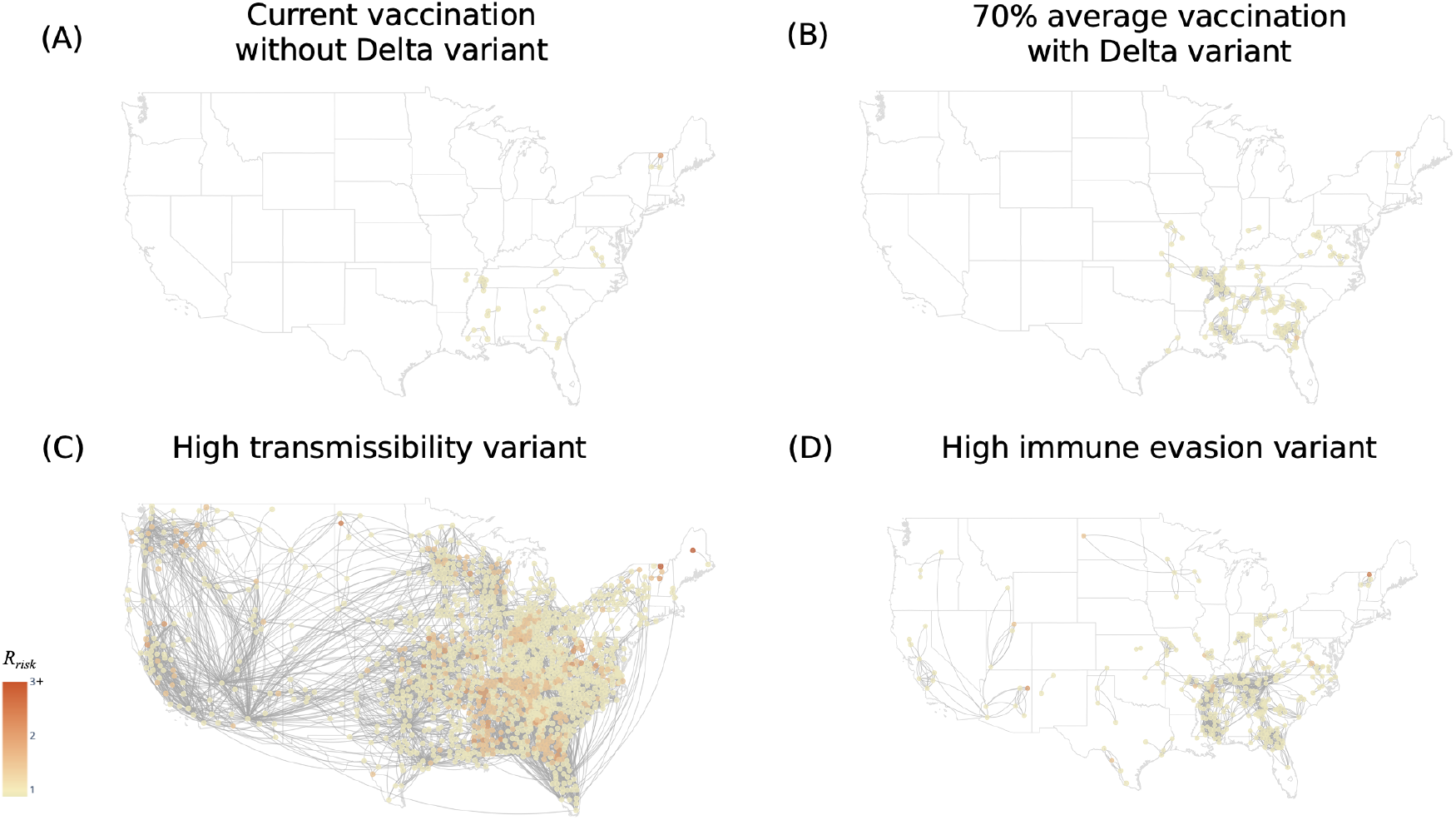
Landscapes of transmission potential vary by scenario. We estimate the connected network of communities that have the potential for *R*^*risk*^*≥* 1 to understand outbreak risk under a number of scenarios. Nodes are county locations with node color denoting *R*^*risk*^, and edges connect locations for which *R*_*ij*_ *>* 0.001. (A) Given current vaccination levels (average US complete vaccination is at ~ 49%) and assuming no introduction of variants, almost all SARS-CoV-2 transmission in the United States would be sub-critical with current levels of contact and mobility. (B) With the Delta variant circulating, a *∼* 70% average vaccination rate would suppress outbreaks in a large part of the country, but some communities would remain vulnerable. (C) Elevated transmissibility (100% increase over wild type) leads to a rise in cases across the majority of counties. (D) High immune evasion (25% decrease in natural resistance/vaccine effectiveness) leads to several disconnected networks in different regions of the country.

## 3 Discussion

Local and regional variability in social mixing as well as heterogeneity in susceptibility are critical in structuring the landscape of COVID-19 transmission potential. In general, the local environment for virus transmission is shaped by both the characteristics of the virus (e.g. transmissibility and virulence) and those of the community it spreads within, such as the effective number of contacts between infected and susceptible individuals. These characteristics vary across space and time based on social dynamics and local public health measures. The conditions in any one community affect not just those within its boundary, but also those linked through mobility. Consequently, realized within-county transmission rates are determined by the local distribution of transmission potential generated by mobility-linked communities – even if any one county has carefully implemented public health measures, individuals within that county are also affected by implementations and compliance in neighboring counties. Therefore, heterogeneity in vaccination rollout and in the relaxation of social distancing and mask mandates will continue to produce heterogeneity in SARS-CoV-2 transmission potential.

While highly effective vaccinations have lead to a decline in incidence in the United States, there remains both current and future SARS-CoV-2 transmission risk. Local risk is affected, but not solely determined, by vaccination rates – vaccination levels high enough to consistently decrease incidence today do not imply that the herd immunity threshold has been permanently crossed. Local immunity can decrease transmission, but does not by itself determine whether *R*_*t*_ *<* 1. Higher contact rates, for example from the return to in-person work or school, or SARS-CoV-2 variants with higher transmissibility or levels of immune evasion can increase *R*_*t*_, potentially causing fresh outbreaks. Movement between counties (and from across the globe) will lead to regular re-introductions of existing and novel variants [20, 21]. Even in settings with low (or sub-critical) effective reproductive numbers, superspreading events can rapidly increase local incidence [22, 23]. Under current conditions we can continue to expect local transmission and patchy outbreaks in areas with low vaccination coverage.

Infectious diseases on the path to local elimination exhibit unique, and often less predictable, dynamics [24, 25]. Fundamental barriers to epidemic forecasting may prevent accurate long-term prediction, especially when below the critical epidemic threshold [26]. Near the critical epidemic threshold, the theory of critical slowing down suggests that stronger surveillance is necessary to keep track of dynamics and that increases in the volatility and autocorrelation of incidence data are leading indicators of resurgent outbreaks [27, 28, 29]. Indeed, disease dynamics can change due to age structure, natural or vaccine-induced immunity, or contact patterns [30, 31]. These host network effects have guided SARS-CoV-2 dynamics throughout the course of the pandemic [32], and may continue to do so during a transition to pathogen endemicity [33].

Our results contribute to the rapidly growing literature on the impact of human mobility on SARS-CoV-2 transmission dynamics. Regional and international travel were crucial to the global dissemination of COVID-19 [34, 35, 36]. Indeed, mobility restrictions have dramatically changed patterns of transmission, decreasing the rate of new infections [34, 37, 21]. Mobility also drives patterns of variant invasion, introducing variants to new locations and enabling their spread across geographic regions [38, 39, 40, 41]. While limiting mobility can be an effective form of epidemic control, inconsistent implementation can be ineffective, with spread between regions likely to continue despite local restrictions [42]. Here, we build on this literature, demonstrating that regional mobility networks drive patterns of COVID-19 transmission throughout the Unites States. Crucially, we identify systematic structure in these networks of transmission potential, with some regions dominated by the dynamics of one metropolitan area, while others are driven by a number of unique local contributions. These dynamics also alter the impact of local public health interventions, like social distancing, and the risk of future surges in cases due to potential variants.

In the United States, vaccination has decelerated transmission rates – decreasing but not eliminating local transmission – yet there remain multiple scenarios in which transmission once again accelerates. Without continued increases in vaccination rates, variants with increased transmissibility present a high risk for successive waves of COVID-19 transmission. These scenarios are neither forecasts nor predictions, rather they illustrate the range of future possibilities and the magnitude of the challenge to public health still presented by the COVID-19 pandemic. In particular, these scenarios demonstrate the perils of the United States’ growing reliance on vaccination as the sole line of defense against COVID-19. A safe return to “normal” levels of social behavior requires high levels of vaccination to prevent transmission. New SARS-CoV-2 variants with higher transmissibility or (partial) immune evasion further increase this burden, potentially making such a strategy infeasible. Under this vaccine-only strategy, heterogeneous county-level vaccination rates are particularly dangerous, with no defense against large outbreaks in clusters of low vaccination. Effective implementation of this vaccine-only strategy will require broader societal acceptance of COVID-19 vaccines, renewed effort to bring vaccinations to those less able to travel or take time off work, and willingness to adapt vaccinations and public health strategy in response to evolving conditions.

This research builds on the outstanding data collection and dissemination by both public and private organizations in response to the immediate crisis of the pandemic. It would not have been possible without their extraordinary effort, but as conditions return to ordinary in many parts of the United States some of these projects are already being halted. Individual-level data collection of this type is vital and must continue over the coming months. Especially when incidence is low, these types of data provide uniquely granular insight into local risk. Averaged quantities on the county, state, or national level can obscure the spatial heterogeneity that we identify as crucial to the dynamics of SARS-CoV-2.

Because we model expected SARS-CoV-2 transmission potential at the county level, our results are limited by our modeling assumptions and the scale of our inference. We assume that county-level seroprevalence deviates from the state-level model in a similar way for all counties. We also assume that response bias in the US CTIS and the CDC seroprevalance survey is multiplicative and constant over time, that the log-odds of case reporting is constant across counties within a given state, and that any seroreversion is negligible. Recent evidence also indicates that the immune response due to natural infection may be less protective against current variants of concern than vaccine-induced immune response [15], which may suggest that our estimates of effective natural protection may be overestimated. Further modeling assumptions (e.g. error distributions, model structures) are discussed in the Supplementary Methods **??, ??**, and **??**. Because the base unit of our model is the county-week, we are restricted to inference on counties. Structure in the network of contact and mixing at the sub-county level or by demographic characteristics such as age could lead to outbreaks even in counties in which we identify this as unlikely. In addition, we model only the first moment of the offspring distribution; we do not model the probability of a superspreading tail-event from the offspring distribution [43, 44].

Despite the declines in incidence in the United States, the COVID-19 pandemic is not over until it is over everywhere. The SARS-CoV-2 virus will continue to evolve and mobility will cause continual reintroductions. Counties with lower rates of immunity remain at elevated risk for outbreaks, especially from new variants. Counties can continue to decrease the risk of these outbreaks by raising vaccination rates and limiting unnecessary contacts, especially in large groups. Indeed, the unvaccinated are at highest risk both of infection and serious illness if infected. Many potential variants are not only more transmissible, but also result in increased severe outcomes [45] – placing the unvaccinated at still higher risk. Over the coming months, continued surveillance of ongoing transmission and of breakthrough cases will be necessary to monitor the potential for more outbreaks (i.e. *R*_*t*_ *>* 1). Although vaccine boosters may at some point become necessary, support of robust local public health capacity can provide an immediate benefit. Effective contact tracing programs and local case surveillance will provide insight into a community’s risk while preventing onward transmission. And although fortifying previously overwhelmed local public health capacity is a vital piece of this effort to end the pandemic. It is also our ethical imperative to increase the global vaccine supply as global vaccination equity will make us all safer. Neither local elimination nor global eradication is feasible in the near-term, but a renewed commitment to public health can dramatically lower local risk, protecting the most vulnerable.

## 4 Methods

We estimate *R*_*ij*_(*t*), the transmission potential produced in community *j* by a case from community *i* at time *t* by combining modeled estimates of contact and seroprevalance and data on mobility and vaccination. From county-specific transmission potential, we generate *R*^*liability*^, the expected number of secondary cases produced anywhere by case in county *i*, and *R*^*risk*^, the expected number of secondary cases in county *j* generated anywhere per case in county *j*. We also account for the impact of variants and residual effects that are not directly modeled from data.

### 4.1 Spatial model of transmission

Using a metapopulation approach, we define (*R*_*ij*_(*t*)) as the number of cases generated in county *j* by a case in location *i* at time *t* (More details about the derivation can be found in the Supplementary Methods **??**). (*R*_*ij*_(*t*)) is a function of the probability a resident of *j* is susceptible (*σ*_*j*_(*t*)), the mobility-weighted interaction rate between counties *i* and *j* (*p*_*ij*_(*t*)), the rate of recovery (*γ*), a location-specific transmissibility rate (*β*_*i*_(*t*)), and a residual effect (*ω*_*j*_(*t*)):

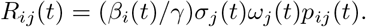

The transmissibility, *β*_*i*_(*t*), is dependent on the relative variant circulation in location *i* at time *t* (see Figure **??**). We parameterize the combination of 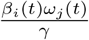 through the residual effect analysis explained below. We adjust this transmissibility to account for variant circulation as explained below.

We estimate the susceptible population size as the proportion of the population with natural immunity (based on the proportion of the population currently infected, *x*_*i*_, and the proportion infected in the past, *r*_*i*_) as well as the proportion with vaccine-derived immunity, based on available estimates on the partial vaccination coverage (i.e. one dose of a two-dose vaccine schedule), 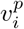, and the complete vaccination coverage, 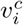. We assume that natural immunity and partial and complete vaccine-induced immunity are imperfect (with an efficacy of *E*_*R*_, *E*_*P*_ and *E*_*C*_, respectively), and we assume that previously infected individuals may be vaccinated. Therefore, the proportion of the population that is still susceptible is determined by:

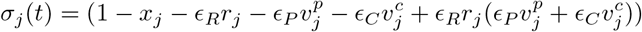

where the final term prevents double counting of individuals with both natural- and vaccine-derived immunity. (More details about the parameterization of *σ*_*j*_(*t*) can be found below).

To capture residual effects stemming from unobserved data or data biases (e.g. systematic differences in contact reporting or in NPIs like mask-wearing), we estimate *ω*_*j*_(*t*) for every location *j* at time *t*. More details on the parameterization of *ω*_*j*_(*t*) can be found below.

We describe the interaction between community *i* and community *j* as the sum of (a) individuals from *i* going to *j* and infecting residents of *j* (b) individuals from *j* coming to *i* and getting infected from residents of *i* and (c) individuals from *i* going to a third community *k* and infecting residents of *j* who are currently in *k*. This estimation assumes that each location has its own household (*α*^*HH*^) and nonhousehold (*α*^*NH*^) contact rate. Additionally, we include an empirically-derived scaling factor (*ζ*_*jt*_) to account for increased transmission risk due to indoor contact. The interaction terms is, therefore, given by:

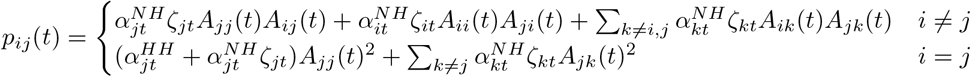

We elaborate on the parameterization of *p*_*ij*_ below.

From *R*_*ij*_(*t*), we can also define:

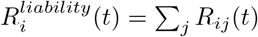: total number of secondary cases generated (anywhere) by a resident of *i*.

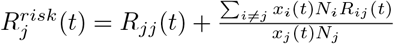is the secondary cases in *j* generated by cases from *j* (*Rjj*(*t*)) and by cases from all *i* (based on incidence (*x*_*i*_(*t*)) and population size (*N*_*i*_) of community *i* at time *t*). We refer to the former as the local component of *R*^*risk*^(*t*) and the latter as the regional component of *R*^*risk*^(*t*).

### 4.2 Natural and vaccine-induced immunity

To parameterize *σ*_*j*_(*t*), we estimate the effective proportion of the population that has protection from natural immunity and vaccine-induced immunity against SARS-CoV-2.

To estimate natural immunity, we use commercial lab SARS-CoV-2 seroprevalence surveys from the Centers for Disease Control and Prevention (CDC), which report roughly biweekly at the state level [46], and county-level weekly SARS-CoV-2 incidence data [47]. To estimate county-level seroprevalence, we weight the CDC’s seroprevalence estimates by county-level cumulative incidence. We assume one state-wide underreporting multiplier *ρ*_*t*_ on the county-level weekly incidence *λ*_*i,t*_ for all counties *i* in week *t*, with the underreporting multiplier *ρ*_*t*_ changing as a linear function of time on the logit scale. This model produces estimates of both county-level seroprevalence and the number of new infections corrected for underreporting in county *i* during week *t*, and we assume that immune response is long-lasting and robust [48, 49, 50]. (More details on the modeling can be found in Supplementary Methods **??**.)

To produce estimates of vaccine-derived immunity, we integrate and correct data from the CDC and state health departments on partial and complete vaccination counts, and we homogeneously normalize these using population estimates from the 2019 American Community Survey [51]. We assume a two week lag for the protective effect of both one-dose and two-dose vaccine-induced immunity. We parameterize vaccine efficacy against circulating variants as described in Supplementary Table **??** [52].

### 4.3 Contact, mobility, and behavioral seasonality

To parameterize *p*_*ij*_(*t*), we use survey data on contacts relevant to respiratory disease transmission, and device-based data on mobility between communities.

To parameterize *α*_*HH*_ and *α*_*NH*_, we aggregate individual-level reported number of non-household epidemiological contacts in the Delphi Group at Carnegie Mellon University’s U.S. COVID-19 Trends and Impact Survey (Delphi US CTIS) to the county-week mean. We model the mean county-level reported contact across time using separate weighted Generalized Additive Models for each state [53], with sample weights applied to the county-week means. In each state-level model, we include the overall state-level trend and county-level deviations from the state-level trend with cubic regression splines and a factor-smooth interaction to share information between counties within the state. A factor-smooth interaction enforces a shared smoothness parameter for all the levels of the factor (i.e. counties within the state) and penalizes the county-specific trends like random effects, allowing the county-specific trend to reduce to the shared state-level trend. We treat household size as constant over the time period of this analysis, deriving a county-specific mean household contact estimate from the survey data. (Additional modeling details can be found in the Supplementary Methods **??**).

To parameterize *A*_*ij*_, we derive county-level estimates of within- and between-county mobility from the Safegraph Social Distancing Metrics dataset [54]. Safegraph uses anonymized mobile app-based GPS location data to capture visits to over 5.5 million points-of-interest in the US daily. We aggregate this visit data to the county- and weekly-scale to produce an estimate of weekly mobility from county *i* to county *j* as the proportion of visits of residents in county *i* that were to a location in county *j* for all *j*. (Additional details can be found in the Supplementary Methods **??**).

Using point-of-interest visit data from the Safegraph Weekly Patterns dataset [55], we also estimate the ratio of mobility occurring at indoor locations relative to outdoor locations which varies seasonally [56] for every county *i* at time *t* and integrate this into the contact estimate *α*_*NH*_ .

### 4.4 Variants

We include the effect of SARS-CoV-2 variants of concern on both transmissibility and vaccine effectiveness. Using data provided by the CDC of genomic sampling at the HHS region level, we construct a mean transmissibility increase in the HHS region at week *t* from reported variant transmissibility relative to wild type weighted by variant genome proportion in the weekly sample. We include the impact of variants on vaccine effectiveness for only the Alpha and Delta variants and produce modified estimates of partial and complete vaccine effectiveness (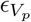 and 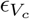) through the same weighted mean procedure (see Table **??**). We assume weekly relative variant proportion is the same for all counties within an HHS region.

### 4.5 Residual effects

Our goal has been to produce a parsimonious generative model to estimate spatio-temporal transmission potential. However, we are unable to integrate data on all disease-relevant processes and the data we have included are subject to bias. Here we seek to integrate these systematic residual effects into our analysis, and we anticipate four key sources of such effects:

1. County-level reporting bias in number of contacts in the US CTIS.
2. County-level reporting bias of local incidence. Our seroprevalence model corrects for this bias with a state-level underreporting multiplier, but systematic county-level deviations from the state mean are not accounted for by the seroprevalance model.
3. State-level bias in the CDC commercial laboratory seroprevalence surveys.
4. Precautionary behaviors such as masking not included in our analysis.

To capture these effects, we estimate a county-specific adjustment factor. Our estimated *R*_*risk,t*_ values are the expectation of the distribution of the effective reproductive number and observed *R*_*t*_ values are autocorrelated realizations of this distribution. Therefore, unbiased *R*_*risk*_ estimates should have the same mean as the observed *R*_*t*_ values. We use this equality to solve for the county-specific factor as 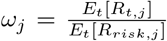 and integrate this factor into the *R*_*ij*_ calculation. (Further details are provided in the Supplementary Methods **??**).

### 4.6 Eigenspectrum analysis

For structured populations, an estimate for the global effective reproductive number can be found using the next-generation matrix method [16, 17]. In our model formulation, *R*_*ij*_ plays the role of the next-generation matrix, and its largest eigenvalue corresponds to the global effective reproductive number, *R*_eff_. To understand global spatial dynamics, we inspect the distribution of eigenvalues, or eigenspectrum, of *R*_*ij*_(*t*). It can be shown that the eigenvalues are all real and non-negative, implying the (unstable) disease-free equilibrium is a saddle point. (Additional details can be found in the Supplementary Methods **??**).

### 4.7 Intervention analysis

To inform the role of different public health interventions on local and regional transmission dynamics, we consider the tradeoff between increasing immunity (e.g. through vaccination) versus reducing contact (e.g. through social distancing). To formalize this, let us consider 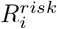 in county *i* as a function of *α*_*j*_ (contacts outside household in county *j*), and 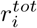 (total estimated population-level immunity in county *i*). Let 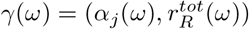 the curve in parameter space, spanned by *ω*, along which 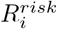 is constant: 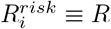. By definition 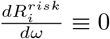 This implies

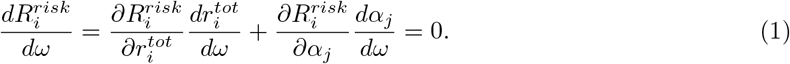

For convenience, we use *α*_*j*_ as the parameter curve: *ω* = *α*_*j*_. From the above expression we can then derive how changes in 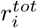 and *α*_*j*_ are linked, through an expression we call *φ*:

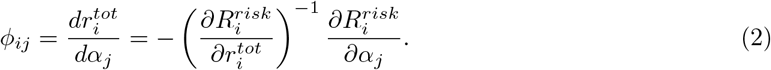

Using the definition of 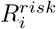 introduced, we get the final expression for *φ*. We also make the time dependence explicit:

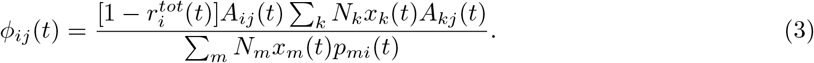

*φ*_*ij*_ estimates what increase in immunity is required in community *i* to achieve the same reduction in *R*^*risk*^ as a decrease in contact in community *j*. We also define the *influencer of location i* as the location for which *φ*_*ij*_ is maximized across all *j*.

### 4.8 Scenario Analysis

To characterize the spatial extent of transmission surges resulting from changes in viral dynamics or public health campaigns, we consider four hypothetical scenarios: (a) current vaccination distribution (with an average of 31.6% complete vaccination rate), natural immunity, contact and mobility (as of June 20, 2021) and only wildtype circulation (i.e. no circulation of variants of concern); (b) 70% average vaccination levels across the country distributed according to current levels of vaccination disparity, and current natural immunity, contact and mobility with current circulating levels of variants of concern (as of June 20, 2021); (c) current levels of vaccination, natural immunity, contact and mobility (as of June 21, 2021) with circulating levels of variants of concern assuming only increased transmissibility compared to wildtype (no immune evasion); and (d) current levels of vaccination, natural immunity, contact and mobility (as of June 21, 2021) with circulating levels of variants of concern assuming only increased immune evasion compared to wildtype (no increase in transmissibility).

For each, scenario, we re-estimate *R*_*ij*_ values assuming the scenario conditions. We then construct the network of nodes (i.e. counties) which have *R*_*risk*_ *>* 1 and edges (i.e. between-county transmission potential) which have *R*_*ij*_ *>* 0.001. (Details on this threshold can be found in the Suppplementary Methods). Thus the resulting network is the set of communities that are at risk for supercritical transmission and have significant capability of introducing infection into the remaining connected communities.

## Data Availability

All data used comes from accessible data repositories. SARS-CoV-2 data is available from the Centers for Disease Control and Prevention (https://covid.cdc.gov/covid-data-tracker/#county-view, https://www.cdc.gov/coronavirus/2019-ncov/cases-updates/geographic-seroprevalence-surveys.html) and the New York Times (https://github.com/nytimes/covid-19-data). Behavior data is available from Safegraph (https://www.safegraph.com/academics) and the Delphi CTIS (https://delphi.cmu.edu/covidcast/surveys/).
All data for the study will be available at https://github.com/bansallab/spatialR

## Acknowledgements

We gratefully acknowledge our data sharing partners. This research is based on survey results from Carnegie Mellon University’s Delphi Group and Facebook; anonymous and aggregated place visit data from Safegraph; incidence data from the New York Times; seroprevalence, vaccination, and variant data from the Centers for Disease Control and Prevention; and vaccination data from state health departments. We thank Alexes Merritt, Andrew Tiu, Benjamin Susswein, Casey Zipfel, Eva Rest, Greg Albery, and Mattia Mazzoli for their contributions and insights. This work was supported by the National Institute of General Medical Sciences of the NIH under Award Number R01GM123007.

