## Supplementary Information for "Ignoring spatial heterogeneity in drivers of SARS-CoV-2 transmission in the US will impede sustained elimination"

July 2021

#### S1 Supplementary Methods

##### S1.1 Contact data and mobility data modeling

We obtain individual-level data on contact rates from the Delphi Group at Carnegie Mellon University U.S. COVID-19 Trends and Impact Survey (CTIS), in partnership with Facebook. This survey is written by the Delphi Group and administered daily to a random sample of Facebook users. The survey defines contact in the question as 'Direct contact' means: "a conversation lasting more than 5 minutes with a person who is closer than 6 feet away from you, or physical contact like hand-shaking, hugging, or kissing." We remove responses with logical inconsistencies (e.g. respondent reports not leaving the house for the past 7 days and going to the grocery store in the past 3 days) and those that report greater than 100 contacts or fewer than 0 contacts over the previous day. After cleaning, we aggregate these individual daily responses to the county-week mean.

We model the county-week means using Generalized Additive Models in mgcv version 1.8-36 [1]. We model the county means across time with a separate model for each state. For each state model, we model a state intercept, a state-level overall trend, and a factor-smooth interaction term. The factor-smooth interaction provides both county-specific random intercepts and county-specific deviations from the overall state trend of time. We fit the state mean trend with a maximum of 30 basis functions and the county-deviations with a maximum of 15 basis functions. The factor-smooth interaction assumes that all the counties have the same "wiggliness" and applies shrinkage on the magnitude of the second derivative of the county-specific deviations. We provide sample weights of the number of responses contributing to the observed county-week mean. We assume that the residuals of the modeled means are normally distributed.

We use mobility data from the Safegraph Social Distancing dataset which provides counts of unique trips between locations, which can be classified as within or between counties. We combine these data into weekly counts of movement between counties and construct a weekly mobility matrix, with each row representing movement from county $i$ to county $j$. We normalize each of these matrices to be row-stochastic, removing elements representing fewer than 5 people or a proportion of mobility smaller then 1e-4. We $z$-score the non-diagonal elements of each week's matrix and clip them between [-3,3].

##### S1.2 County-level natural immunity rates

We model county-level rates of natural immunity using state-level seroprevalance estimates from the CDC's Commercial Laboratory Seroprevalence Surveys for COVID-19. This program partners with commercial laboratories processing routine non-COVID-19 related blood samples to produce roughly biweekly estimates state populations' COVID-19 seroprevalence. The program's methodology is explained in [2].

From the state seroprevalance estimate, we construct county-level estimates by stratifying by reported cumulative incidence using incidence data from The New York Times, based on reports from state and local health agencies. We assume one statewide underreporting multiplier, $\rho_{A,t}$, on the county-level weekly incidence $\lambda_{i,t}$ in a state $A$ for all counties $i$ in week $t$, with the underreporting multiplier $\rho_{A,t}$ changing as a linear function of time on the logit scale. We model the seroprevalence in state $A$ on week $t$ as binomially distributed, with the number of seropositives , $N_{pos,A,t}$ and total samples $N_{sample,A,t}$ the results from the CDC survey in that state-week. We model the observed weekly incidence on week $t$ in county $i$ as Poisson distributed. This Poisson-distributed rate is scaled to correct for underreporting on the logit scale, with one state-level multiplier applied to every county within the state. On the logit scale, the underreporting multiplier $\rho_t$ is a simple linear function of time. The proportion of the population seropositive $p_{seroconverted}$ is a lagged version of the cumulative weekly incidence in order to account for time to seroconvert post-infection [3].

$$p_{pos,A,t} \sim Bin(N_{sample,A,t}, N_{pos,A,t})$$

$$p_{pos,A,t} = \sum_{\tau=0}^{t} (p_{seroconverted,A,\tau})$$

$$p_{seroconverted,A,t} = 0.25p_{weekly,A,t-1} + 0.5p_{weekly,A,t-2} + 0.25p_{weekly,A,t-3}$$

$$p_{weekly,A,t} = \sum_{i} p_{weekly,i,t}, \forall i \in \{\text{State A}\}$$

$$logit(p_{weekly,i,t}) = \rho_t + logit(\frac{\lambda_{i,t}}{Pop_i})$$

$$\rho_t \sim N(\alpha_0 + \alpha_1(t/t_{max}), \sigma_\rho^2)$$

$$Incidence_{i,t} \sim Pois(\lambda_{i,t})$$

The priors for model parameters are:

$$\lambda \sim [0, \infty]$$

$$\alpha_0 \sim N(2.5, .15)$$

$$\alpha_1 \sim N(0, 1)$$

$$\sigma_\rho^2 \sim t(3, 0, 1)$$

This model produces estimates of both county-level seroprevalence and the number of new infections corrected for underreporting in county $i$ during week $t$. We fit the model in CmdStanR version 0.4.0.9000 for 2000 warmup iterations and 2000 sampling iterations for 4 chains [4]. All split rhat diagnostics are below 1.01, indicating model convergence.

Due to irregularities in the CDC data in the states of North Dokota and New York, we adjust our modeling strategy for these states. In North Dakota, we take antibody testing data from the COVID Tracking Project and adjust for careseeking bias with a binomial regression model by regressing the CDC antibody data on the COVID Tracking Project antibody test positivity rates in brms version 2.15.0 [5]. We apply the fitted values from this model as the population positivity rates in the seroprevalance model. For New York, we do not fit a model and instead apply the posterior estimates of the underreporting multiplier for New Jersey to produce county seroprevalance estimates.

This model relies on a number of assumptions. It assumes if there is COVID-19 circulating in the community that there is at least some non-zero number of cases reported in order to be scaled for underreporting. It cannot correct for the complete absence of case reporting. There is only one state-level multiplier, so any heterogeneity in underreporting across counties is ignored (but see the Residual effect analysis subsection). The model assumes that underreporting changes as a linear function of time on the logit scale. The model assumes that the proportion of the county seropositive in any given county is not very close to 1. We assume that any antibody waning in negligible. We check for model capability by simulating from and recovering parameters.

Using these estimates and data from the CDC and state Departments of Health on vaccination rates, we produce estimates of of the number of COVID-19 immune individuals in each county for a given week as described in Subsubsection 4.1. We assume a two week lag for the protective effect of both one-dose and two-dose vaccine-induced immunity.

#### S1.3 Residual effect analysis

We generate county-level estimates of residual effects not accounted for by our mechanistic modeling approach. In the mechanistic component of our model, we combine contact, mobility, local immunity, and local incidence to generate our $R_j^{risk}$ estimates, as detailed in the "Metapopulation model" subsubsection. These $R_j^{risk}$ values are modeled estimates of the mean of $R_t$ in county $j$. In other words, they are estimates of the conditional mean – the first moment – of the offspring distribution for a county-week. Within a county, observed $R_t$ values are autocorrelated draws from this offspring distribution. We do not model the second moment of the offspring distribution and so do not produce estimates of the variance of $R_t$ values around our estimated distributional means, the $R_j^{risk}$.

We anticipate our residual effects accounting for several different sources of variation. While we mechanistically account for social distancing, we do not generatively model other NPIs like mask wearing in the main analysis — we anticipate that these county-specific residual effects can partially account for NPIs like mask wearing. Indeed, in S5, we demonstrate that the residual effects are correlated with proportion reporting mask wearing at the county level (Pearson's rho = 0.15). We also anticipate that systematic response bias in contact number in the CTIS, systematic sampling bias in the CDC commercial laboratory seroprevalence surveys, and incidence underrereporting relative to the state underreporting multiplier could lead to biased county-level $R_j^{risk}$ estimates. Because $R_{ij}$ estimates are the product of these effects, we anticipate that the resulting form of any unmodeled residual effect would be multiplicative in nature.

Because the $R_j^{risk}$ values are estimates of the mean of $R_t$, these two quantities should have the same mean. Assuming that the form of these biases and unaccounted for effects are multiplicative with respect to $R_j^{risk}$ and constant over time, we can estimate the the residual effects by comparing the mean of the $R_j^{risk}$ estimates to that of empirical measurements of $R_t$:

$$\frac{E_t[R_{t,j}]}{E_t[R_{risk,j}]} = \frac{\beta_j \omega_j}{\gamma}$$

where $\beta_j$ is the transmissibility of COVID-19 with the county $j$-specific multiplicative residual effect and $\gamma$ is the COVID-19 recovery rate.

We estimate the $R_j^{risk}$ values as specified in the methods subsection. We estimate $R_t$ at the county-level using the R package EpiEstim version 2.2-4 [6]. We base these estimates on incidence data reported by the New York Times, as described in the Methods subsection. We use this incidence data to generate values of $R_t$ from EpiEstim using a parametric serial interval with a specified mean of 5.5 and standard deviation of 4.5, consistent with [7, 8, 9]. We estimate $R_t$ with a 7-day sliding window.

#### S1.4 Scenario Analysis

In the scenario analysis, we examine the network of connected counties experiencing case growth under the specified conditions. We define counties as "connected" if the (directed) edge weight between the two is greater than 0.001. We select this cutoff using the empirical $R_{ij}(t)$ matrix in Michigan in mid-March; at this point in time there was a surge in COVID-19 cases that spread rapidly throughout the state of Michigan, but this surge did not immediately spread across state borders. We apply this empirical phenomenon to select our cutoff for connected counties in our scenario network: most Michigan within-state edge weights are above 0.001, but almost all between-state edge weights are less than 0.001.

We generate these scenarios using the empirical contact, mobility, vaccination, seroprevalance, and variant data the weeks of May 16, 2021 through June 20, 2021. We estimate the $R_{ij}$ edge weights and $R_j^{risk}$ values for each of these weeks and generate the presented values by averaging over these weeks to decrease stochasticity in the scenario estimates.

#### S1.5 Eigenspectrum Analysis

To understand the spatial dynamics, we analysed the eigenvalues of the reproductive matrix, $R(t)$ (with elements $[R(t)]_{ij} = R_{ij}(t)$). Interpretation of the eigenvalues of $R$ can be aided by recognising that they that they are all non-negative real numbers. The rest of this section is focused on demonstrating this.

For convenience we recapitulate the definitions of $R_{ij}(t)$ and $p_{ij}(t)$,

$$R_{ij}(t) = (\beta_i(t)/\gamma)\sigma_j(t)\omega_j(t)p_{ij}(t), \tag{1}$$

and

$$p_{ij}(t) = \begin{cases} \alpha_{jt}^{NH} A_{jj}(t)A_{ij}(t) + \alpha_{it}^{NH} A_{ii}(t)A_{ji}(t) + \sum_{k\neq i,j} \alpha_{kt}^{NH} A_{ik}(t)A_{jk}(t) & i \neq j, \\ \alpha_{jt}^{HH+NH} A_{jj}(t)^2 + \sum_{k\neq j} \alpha_{kt}^{NH} A_{jk}(t)^2 & i = j. \end{cases} \tag{2}$$

The remaining symbols are defined in the main text. Using Eq. 1, we can write the reproductive matrix $R(t)$ as the matrix product

$$R(t) = Q(t)P(t)S(t) \tag{3}$$

where $P(t)$ has elements $[P(t)]_{ij} = p_{ij}$ and both $Q(t)$ and $S(t)$ are diagonal matrices with elements $[Q(t)]_{ij} = (\beta_i(t)/\gamma)\delta_{ij}$ and $[S(t)]_{ij} = \sigma_j(t)\omega_j(t)\delta_{ij}$ respectively (with $\delta_{ij} = 1$ if $i = j$ and 0 otherwise).

We can substantially simplify the expression for $p_{i,j}$ by defining $b_{ik}(t) = \sqrt{\alpha_{kt}^{NH}} A_{ik}(t)$ and $c_j(t) = (\alpha_{jt}^{HH+NH} - \alpha_{jt}^{NH})A_{jj}(t)^2$, to give

$$p_{ij}(t) = \sum_k b_{ik}(t)b_{jk}(t) + \delta_{i,j}c_j(t). \tag{4}$$

In matrix notation,

$$P(t) = B(t)B(t)^T + C(t) \tag{5}$$

where the matrix $B(t)$ has elements $[B(t)]_{ij} = b_{ij}(t)$ and $C(t)$ is a diagonal matrix with elements $[C(t)]_{ij} = c_j(t)\delta_{ij}$. As can be seen by exchanging $i$ and $j$ indices in Eq. 4, the matrix $P(t)$ is the sum of two symmetric matrices, and is therefore itself symmetric.

While $P(t)$, $Q(t)$ and $S(t)$ are all symmetric, their product is in general not. We can however prove that the eigenvalues of $R(t)$ are real and non-negative. In what follows we will make use of the matrix square root, $X = X^{1/2}X^{1/2}$. For diagonal matrices this is trivially $[X^{1/2}]_{ij} = x_{ij}^{1/2}\delta_{ij}$. The inverse matrix $X^{-1/2}$ has elements $[X^{-1/2}]_{ij} = x_{ij}^{-1/2}\delta_{ij}$.

We begin by defining the matrix $R'$ via the similarity transformation,

$$R' = M^{-1}RM, \tag{6}$$

where the matrix $M = Q^{1/2}S^{-1/2}$. Substituting in the definitions of $M$ and $R$, we find (after some matrix algebra),

$$R' = S^{1/2}Q^{1/2}PQ^{1/2}S^{1/2}. \tag{7}$$

It is straight forward to show this matrix is symmetric. Furthermore, by defining $G = S^{1/2}Q^{1/2}(B+C^{1/2})$ and using Eq. 5, we see that

$$R' = GG^T. \tag{8}$$

The matrix $R'$ is therefore positive semi-definite, as for any arbitrary vector $x$, $x^T R' x = |G^T x|^2 \geq 0$ (where $|y|^2$ is the Euclidean norm of $y$). Since the matrices $R'$ and $R$ are related via a similarity transformation, they share the same eigenvalues. Positive semi-definite matrices have non-negative real eigenvalues, therefore $R$ also has non-negative real eigenvalues.

### S2 Supplementary Figures

| | partial vaccination efficacy | complete vaccination efficacy |
|---|---|---|
| Wildtype | 0.6 | 0.9 |
| Alpha variant | 0.5 | 0.9 |
| Delta variant | 0.33 | 0.9 |

Table S1: Partial and complete vaccination efficacy against circulating variants.

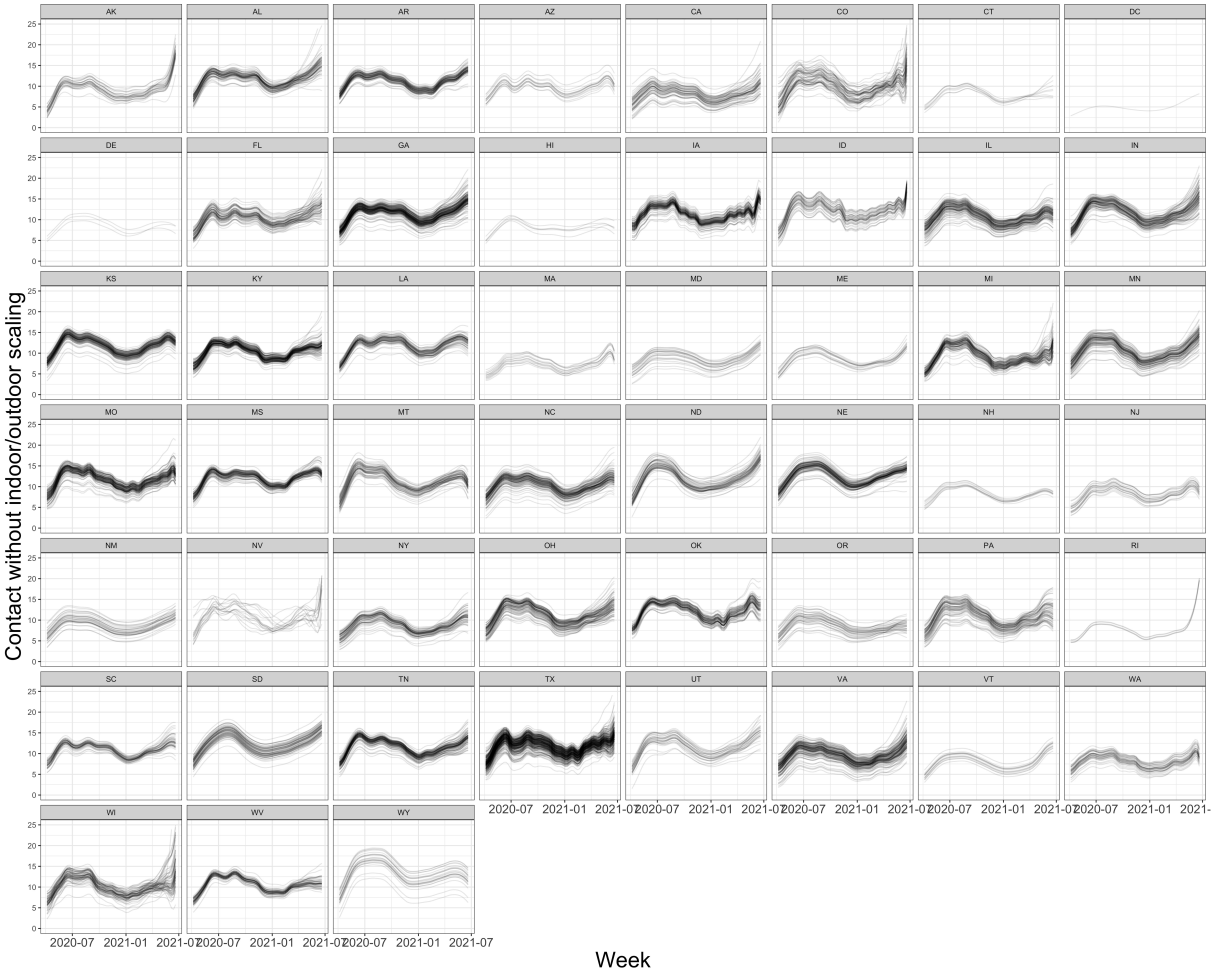


Figure S1: **County-specific contact rates without indoor/outdoor scaling in each state across time.**

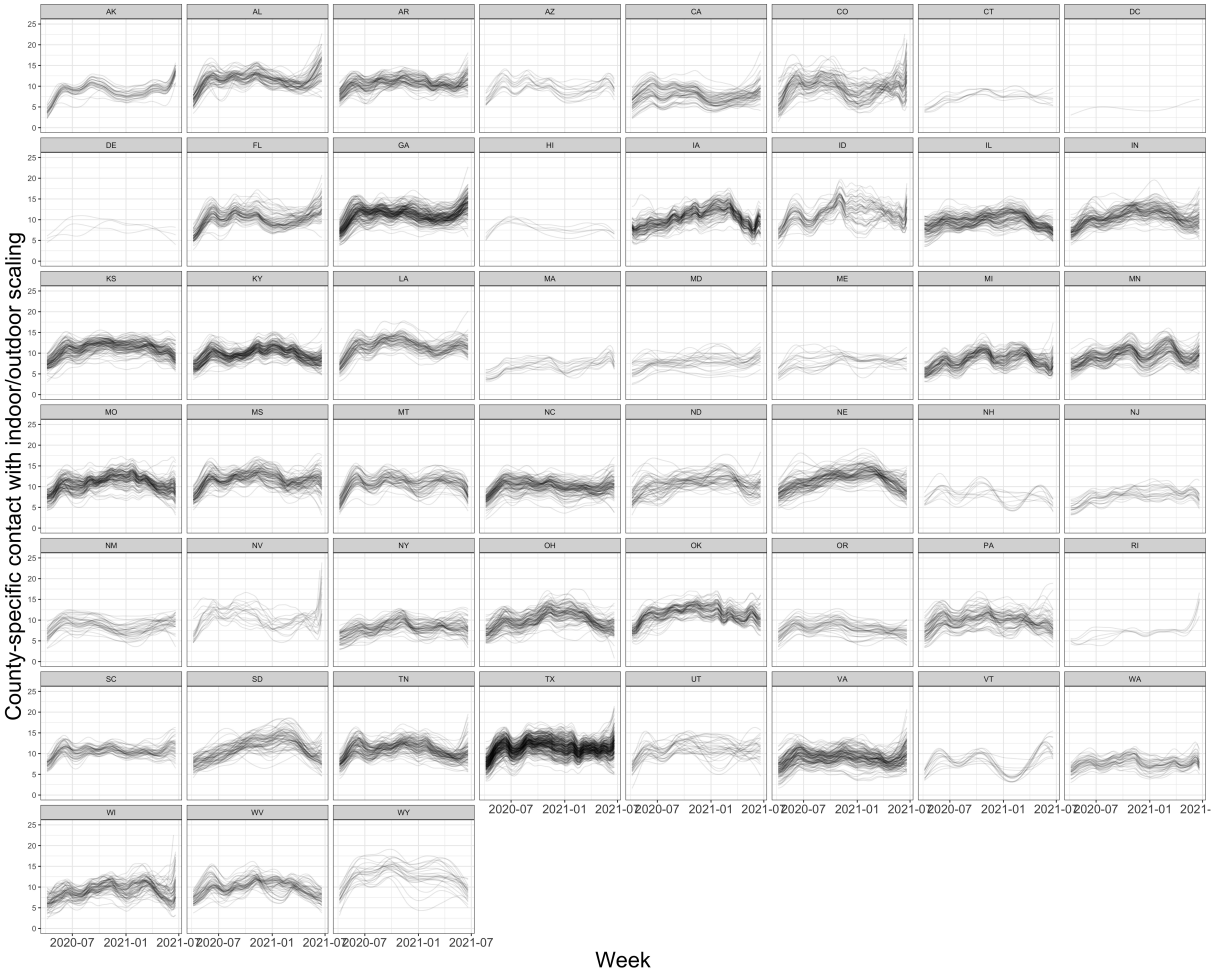


Figure S2: **County-specific contact rates with indoor/outdoor scaling in each state across time.**

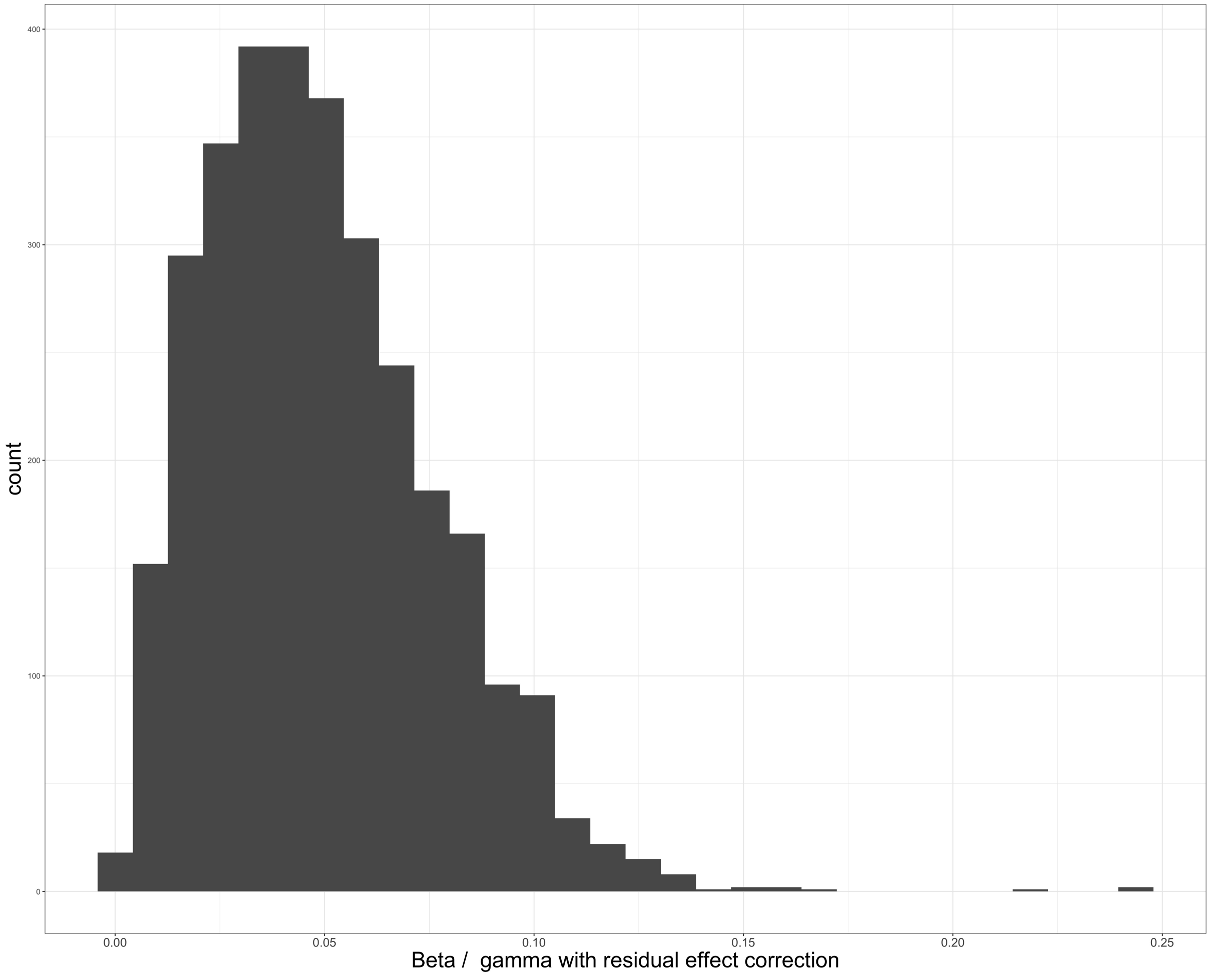


Figure S3: $\beta_i \omega_j / \gamma$ **values** The presented estimates of these $\beta_i \omega_j / \gamma$ values are for pre-variant introduction in the United States and assume wild type transmissibility.

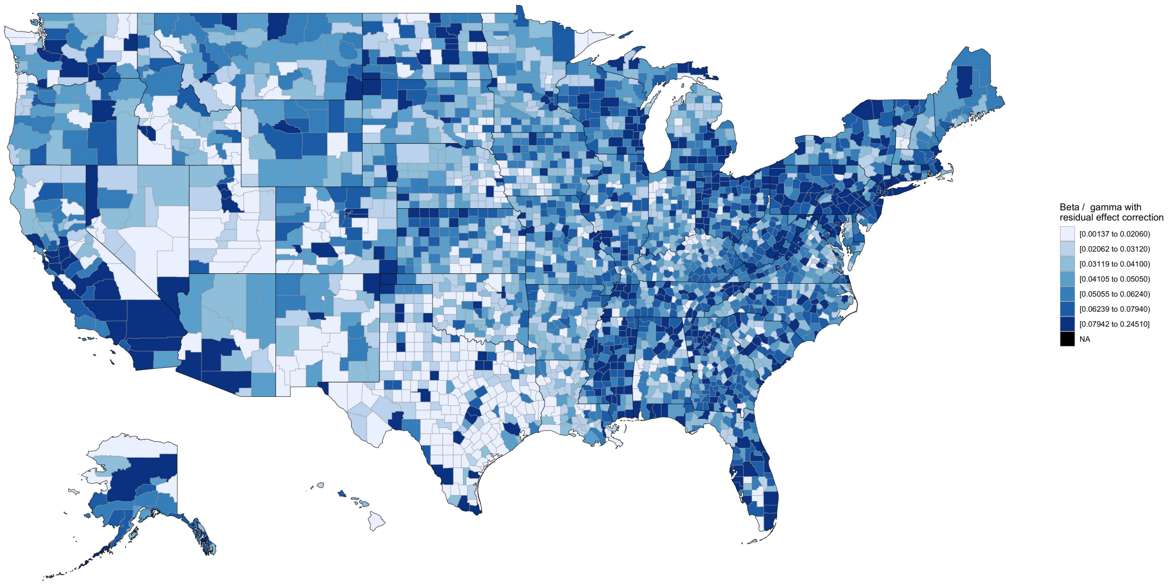


Figure S4: **Choropleth of $\beta_i\omega_j\gamma$ values** The presented estimates of these $\beta_i\omega_j/\gamma$ values are for pre-variant introduction in the United States and assume wild type transmissibility.

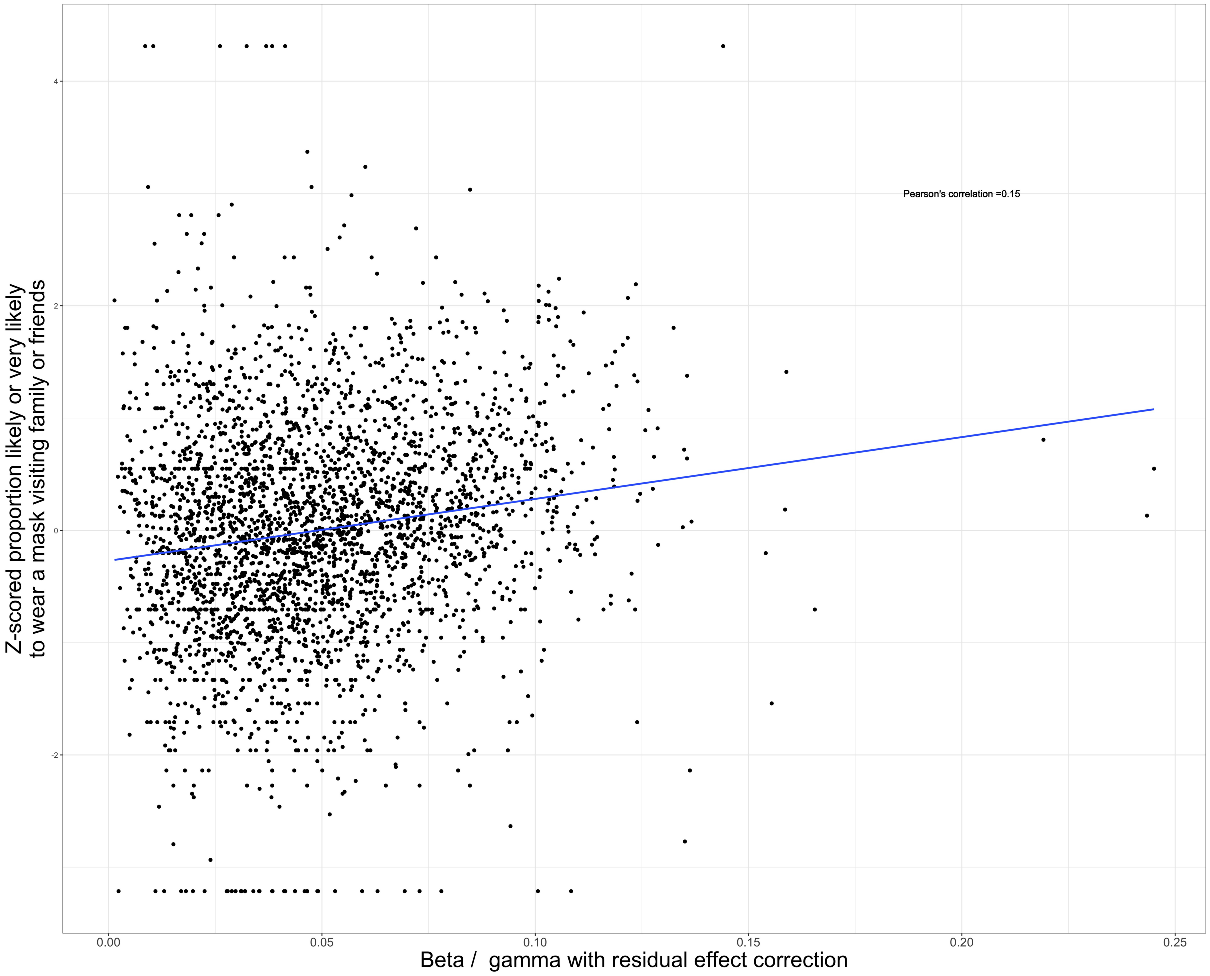


Figure S5: $\beta_i \omega_j / \gamma$ **values are weakly correlated with mask wearing** The presented estimates of these $\beta_i \omega_j / \gamma$ values are for pre-variant introduction in the United States and assume wild type transmissibility.

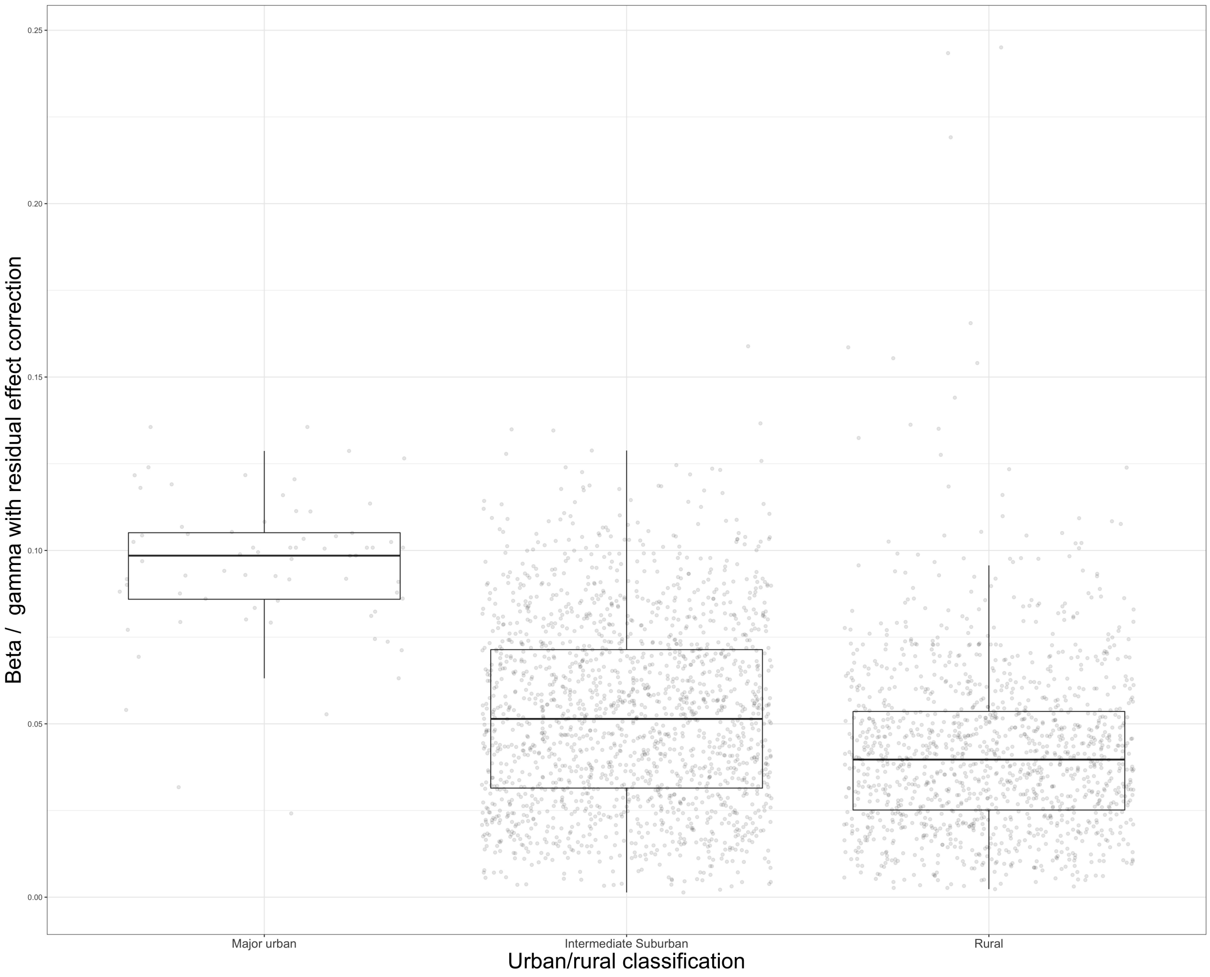


Figure S6: $\beta_i\omega_j/\gamma$ **values vary across urban/rural classification** The presented estimates of these $\beta_i\omega_j/\gamma$ values are for pre-variant introduction in the United States and assume wild type transmissibility.

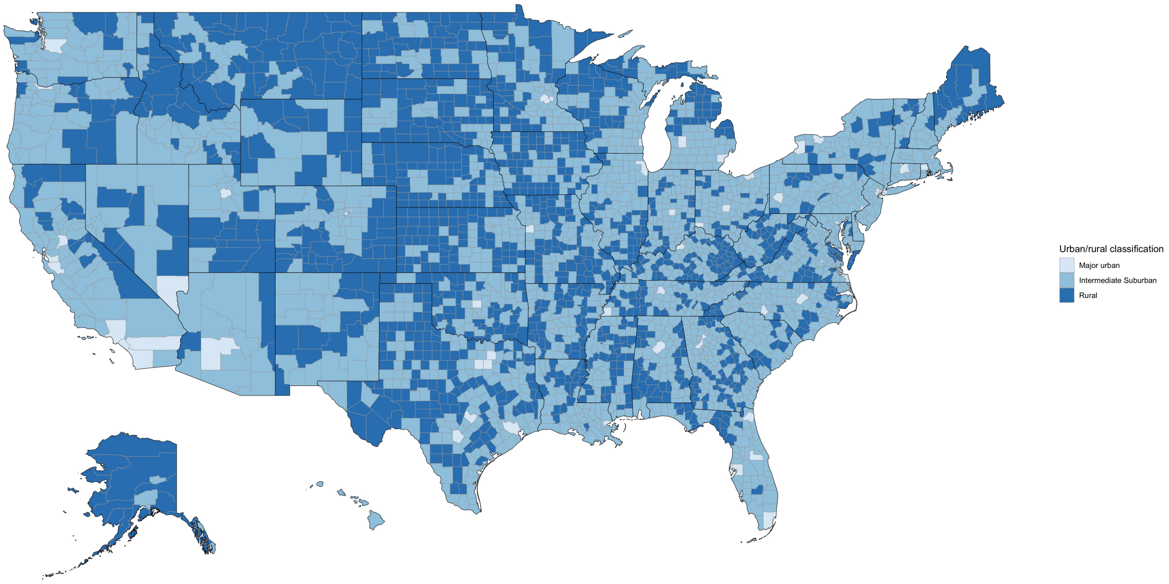


Figure S7: **Choropleth of urban/rural classification**

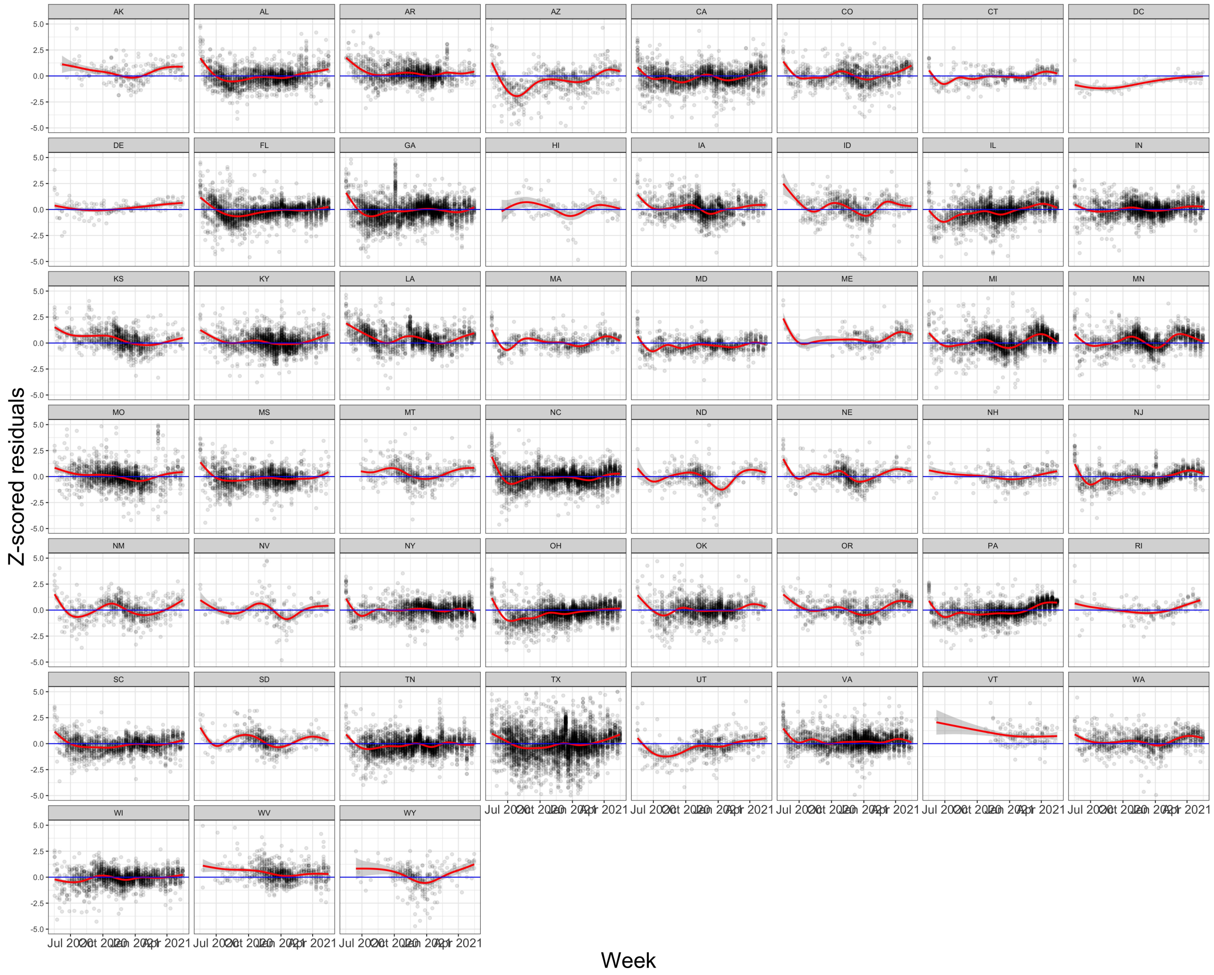


Figure S8: **County-week residuals demonstrate no remaining trend across time**

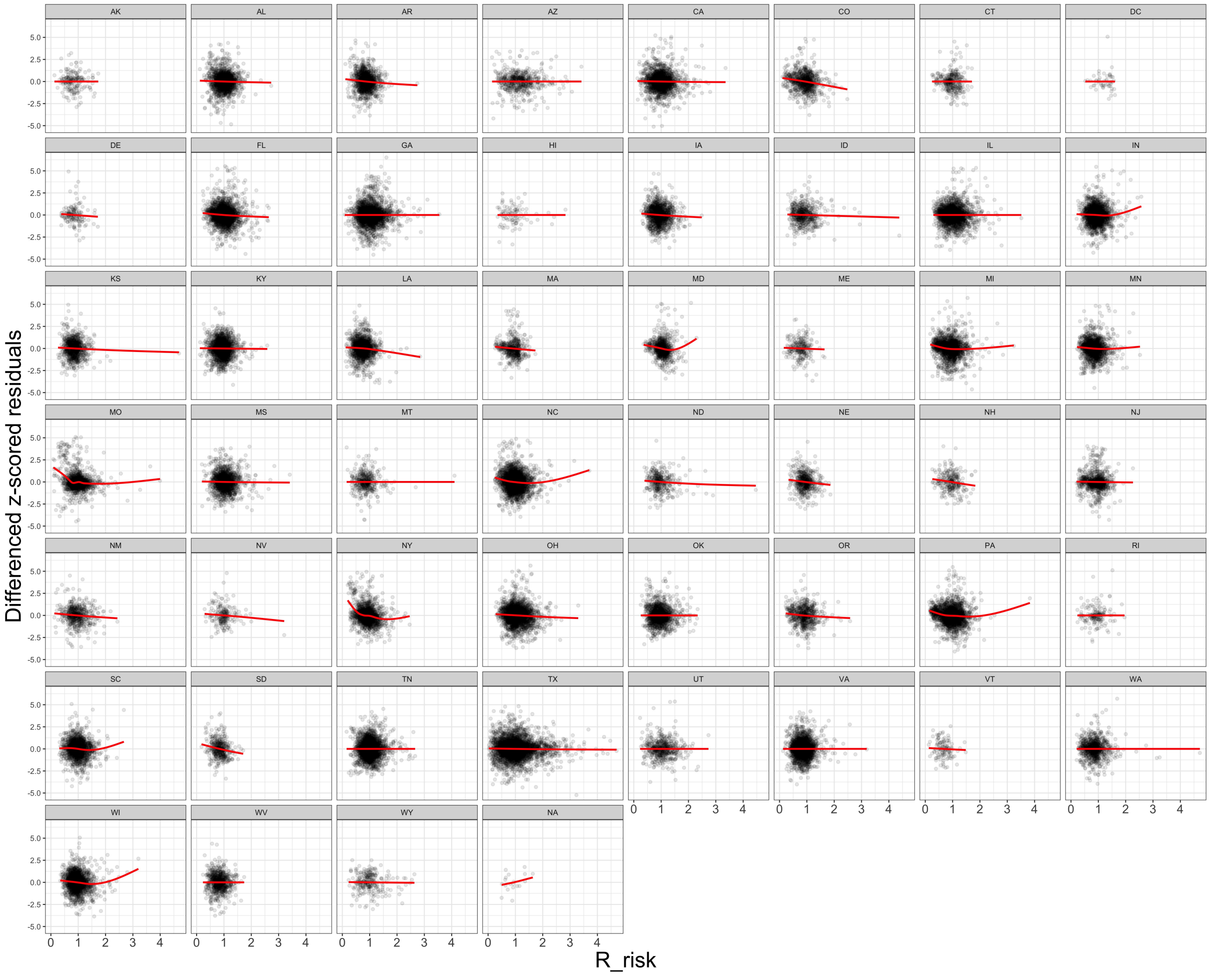


Figure S9: **Differenced county-week residuals demonstrate no remaining trend against $R^{risk}$ values**. Note that we use differenced residuals because the $R^{risk}$ values are estimates of the mean of the distribution, while $R_t$ values are autocorrelated draws from this distribution because of correlation in the underlying incidence data. We do not model this autocorrelation and therefore difference the residuals to look for remaining, non-temporal trend.

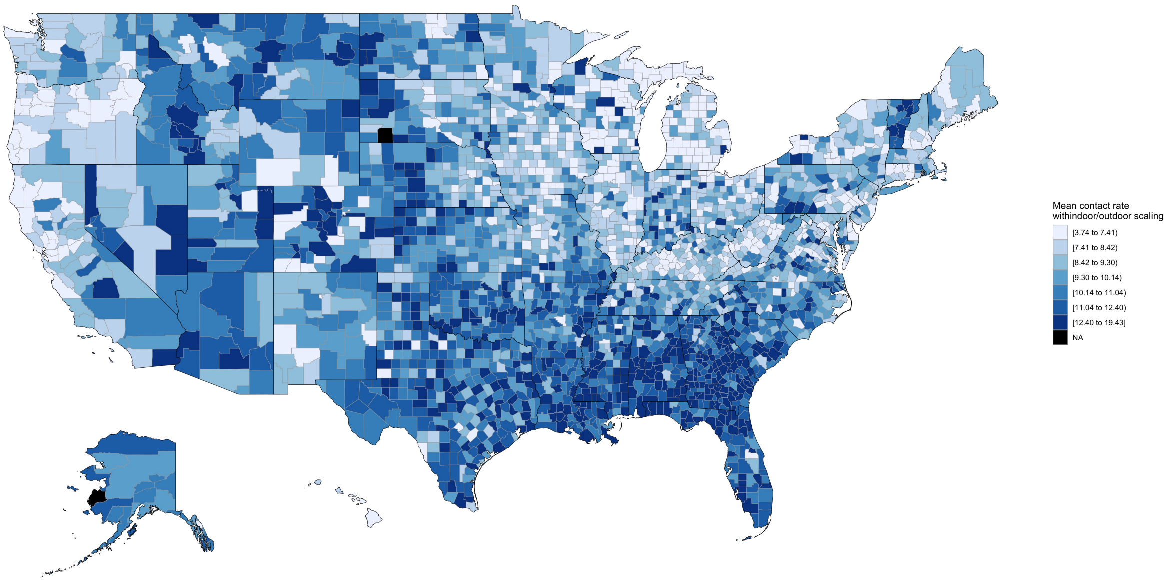


Figure S10: **Choropleth of mean contact rate with indoor/outdoor scaling during scenario time range** The scenarios use the mean of the estimated for the weeks between May 16, 2021 and June 20, 2021.

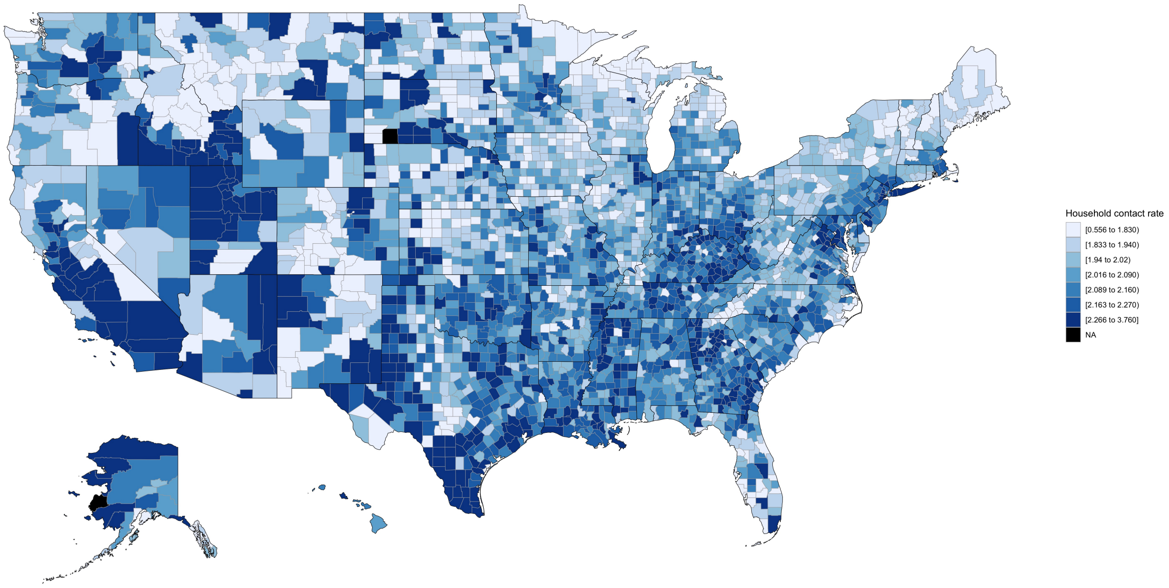


Figure S11: **Choropleth of mean proportion immune during scenario time range** The scenarios use the mean of the estimated for the weeks between May 16, 2021 and June 20, 2021.

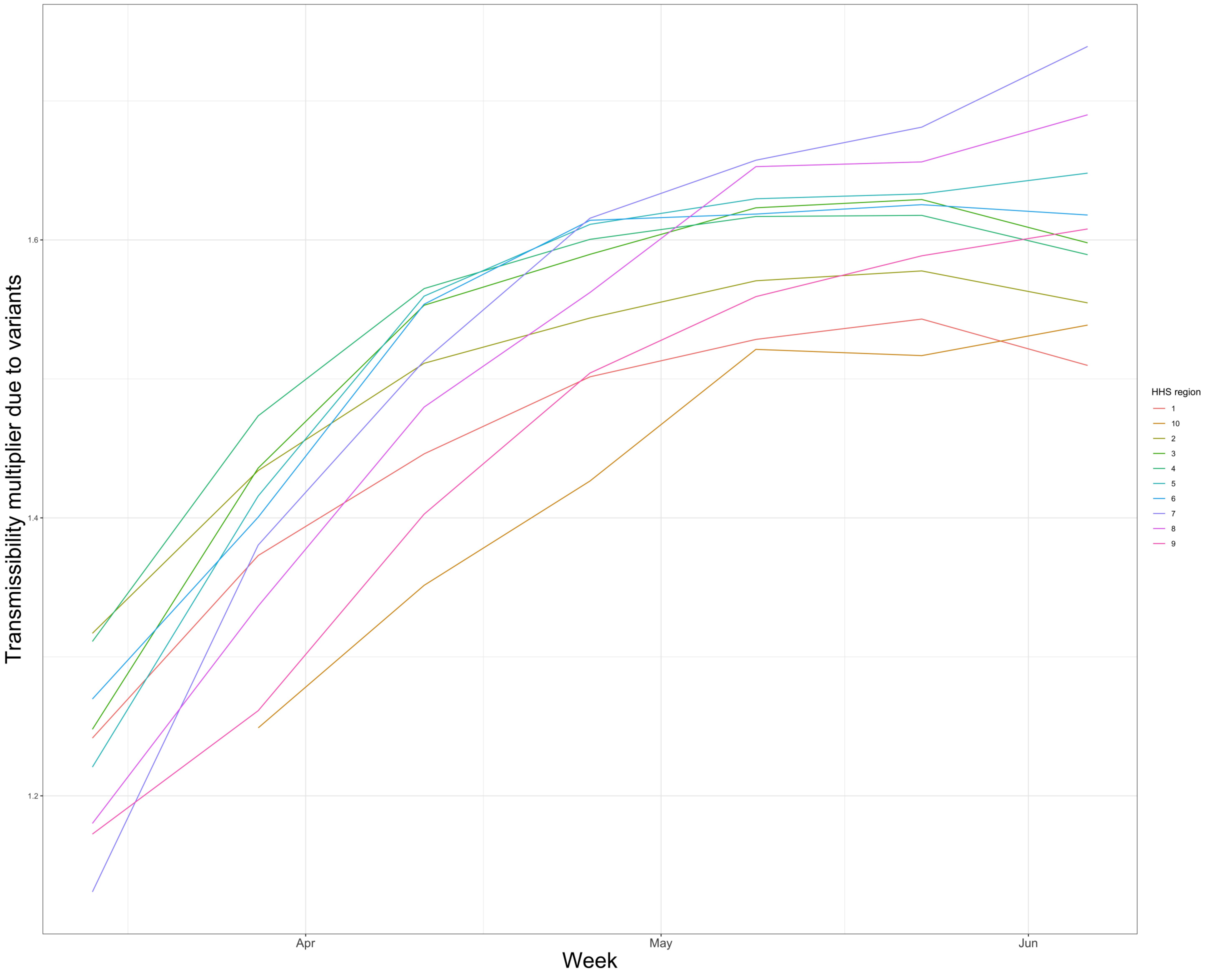


Figure S12: **Transmissibility multiplier due to variant prevalence over time by HHS region**
